# Regional blood flow signatures of opioidergic modulation of ketamine in major depressive disorder: a randomised crossover study

**DOI:** 10.1101/2025.08.18.25333599

**Authors:** Luke A. Jelen, Owen O’Daly, Fernando O. Zelaya, James M. Stone, Allan H. Young, Mitul A. Mehta

**Author notes:** Corresponding author Institute of Psychiatry Psychology & Neuroscience, King’s College London, 16 De Crespigny Park, London SE5 8AF, United Kingdom.

## Abstract

**Objective:** Accumulating evidence suggests the opioid system may modulate ketamine’s rapid antidepressant effects. The objective of this study was to test whether opioid system modulation via naltrexone alters ketamine’s acute effects on regional cerebral blood flow (rCBF) in major depressive disorder (MDD), and whether these changes relate to symptom measures and map onto receptor density profiles.

**Methods:** In a randomised, double-blind, crossover study, 26 adults (18–50 years) with MDD completed two sessions: oral naltrexone 50 mg or placebo, each followed by intravenous ketamine 0.5 mg/kg over 40 minutes during 3D pseudo-continuous arterial spin labelling (3D-pCASL) MRI to quantify rCBF. Subjective effects were assessed with the Clinician-Administered Dissociative States Scale and the Psychotomimetic States Inventory; clinical outcomes with the Montgomery–Åsberg Depression Rating Scale and the Quick Inventory of Depressive Symptomatology Self-Report. Exploratory analyses spatially correlated CBF maps with receptor density profiles (MOR, KOR, NMDA, mGluR5, GABAA, GABAAα5), correcting for spatial autocorrelation.

**Results:** Ketamine increased CBF in subgenual, pregenual, and dorsal anterior cingulate cortices (p < 0.05, voxel-wise FWE-corrected), effects not attenuated by naltrexone. Under placebo pretreatment, baseline-adjusted infusion pregenual relative rCBF was associated with acute subjective effects (PSI-delusional: r = 0.56, p = 0.004; PSI-perceptual distortion: r = 0.64, p < 0.001), and baseline subgenual rCBF (adjusted for global CBF) was associated with day-one antidepressant response (MADRS r = 0.60, p = 0.002; QIDS-SR r = 0.67, p < 0.001). Naltrexone pretreatment disrupted these associations. Ketamine-induced CBF changes aligned with MOR and mGluR5 receptor profiles; naltrexone’s interaction aligned with MOR, mGluR5 and GABAAα5 (pSA-corr < 0.05).

**Conclusions:** This study suggests that ketamine’s effects on CBF in MDD are influenced by complex interactions between glutamatergic, opioidergic, and GABAergic systems. These findings provide mechanistic insights with potential implications for optimising ketamine-based treatments.

**Registration:** ClinicalTrials.gov Identifier: NCT04977674

## Introduction

At subanaesthetic doses, ketamine can produce rapid reductions in depressive symptoms and suicidal ideation, including in individuals with treatment-resistant depression (TRD) (1). Neuroimaging studies have explored ketamine’s effects on brain structure, function, connectivity, neurochemistry, and metabolism, aiming to elucidate its therapeutic mechanisms (2). Most of the functional studies have employed blood oxygen-level-dependent (BOLD) MRI, using both resting-state and task-based paradigms (2). These studies consistently report alterations in brain activity and network functioning, but the relationship between these changes and symptom improvement, as well as the contributions of underlying neurotransmitter systems, remains unclear.

In addition to its primary action as an uncompetitive NMDA receptor antagonist, ketamine also modulates the opioid system (3,4). Preclinical and clinical evidence suggests that opioid system activation contributes to ketamine’s rapid antidepressant effects (4–10). However, this relationship is not consistently observed across all studies (11,12). Understanding how opioid mechanisms influence ketamine’s effects on brain activity and their relationship to clinical outcomes is crucial for clarifying its mechanism of action.

One fundamental aspect of ketamine’s neurovascular effects is its impact on regional cerebral blood flow (rCBF), which may be mediated by both neurovascular coupling and direct vascular effects (13). While BOLD imaging indirectly reflects changes in brain activity through changes in CBF and in cerebral metabolic rate of oxygen consumption (CMRO_2_), arterial spin labelling (ASL) provides direct, quantitative measures of rCBF without ionising radiation or contrast agents (14). ASL has been used to examine the acute effects of psychoactive compounds, including ketamine (15), as well as opioid agonists and antagonists (16).

ASL and PET studies consistently show ketamine-induced rCBF increases in prefrontal, orbitofrontal, anterior cingulate, insular, and subcortical regions including the thalamus, caudate, putamen, and hippocampus (15,17–20). However, the relationship between these rCBF changes and antidepressant response is less consistent. Prior ASL studies in TRD patients have reported variable rCBF alterations following ketamine, with some studies associating these changes with clinical improvement (21–23).

While the effects of ketamine on rCBF have been characterised during intravenous (IV) administration in healthy individuals (15,17), no published studies have examined these effects in MDD patients during acute IV administration. Furthermore, the impact of opioid antagonism on ketamine-induced rCBF changes has not been tested in any population.

In this randomised, double-blind, crossover study, participants with MDD received two separate IV ketamine infusions during neuroimaging, each preceded by either the opioid antagonist naltrexone or placebo. Our primary neuroimaging outcome, previously reported (4), demonstrated that naltrexone attenuated the acute ketamine-induced increase in glutamate+glutamine (Glx) to total N-acetylaspartate (tNAA) ratio and the reduction in Montgomery-Åsberg Depression Rating Scale (MADRS) scores on day 1 post-infusion. Here, we focus on the ASL outcomes, examining how naltrexone modulated ketamine’s effects on rCBF. We hypothesised that ketamine would increase rCBF in regions implicated in emotional processing, including the subgenual, pregenual, and dorsal anterior cingulate cortex (sgACC, pgACC, dACC), as well as the thalamus, and that naltrexone would attenuate this effect. Additionally, we explored the spatial relationship between ketamine-induced rCBF changes and receptor density profiles to better understand the involvement of underlying neurotransmitter systems.

## Methods

### Study Design and Procedures

Ethical approval was obtained from the London – City & East Research Ethics Committee (Reference: 21/LO/0334) and all participants provided written informed consent (ClinicalTrials.gov registration: NCT04977674).

Participants completed two study visits, each involving a double-blind oral pretreatment (naltrexone 50 mg or placebo, ascorbic acid 50 mg), administered 1 hour before a 40-minute IV ketamine infusion (0.5 mg/kg) during MRI. All scans took place at midday for all participants and sessions. The two visits were separated by a 2–4-week washout period (**Figure 1A**). Pretreatment order was randomised 1:1 using a computer-generated block scheme (block size: 4) stratified by sex, resulting in balanced allocation (Placebo-Naltrexone: N = 13; Naltrexone-Placebo: N = 13). Pharmacy staff masked the identity of treatments by over-encapsulating both placebo and naltrexone pills. During MRI scanning, participants were continuously monitored by a study physician, with real-time assessment of respiration, pulse oximetry, and vital signs to ensure safety throughout the infusion. 3D pseudo-Continuous Arterial Spin Labelling (3D-pCASL) data were acquired twice per visit: at baseline (pre-ketamine administration) and during the final 10 minutes of the ketamine infusion (**Figure 1B**).

**Figure 1:**
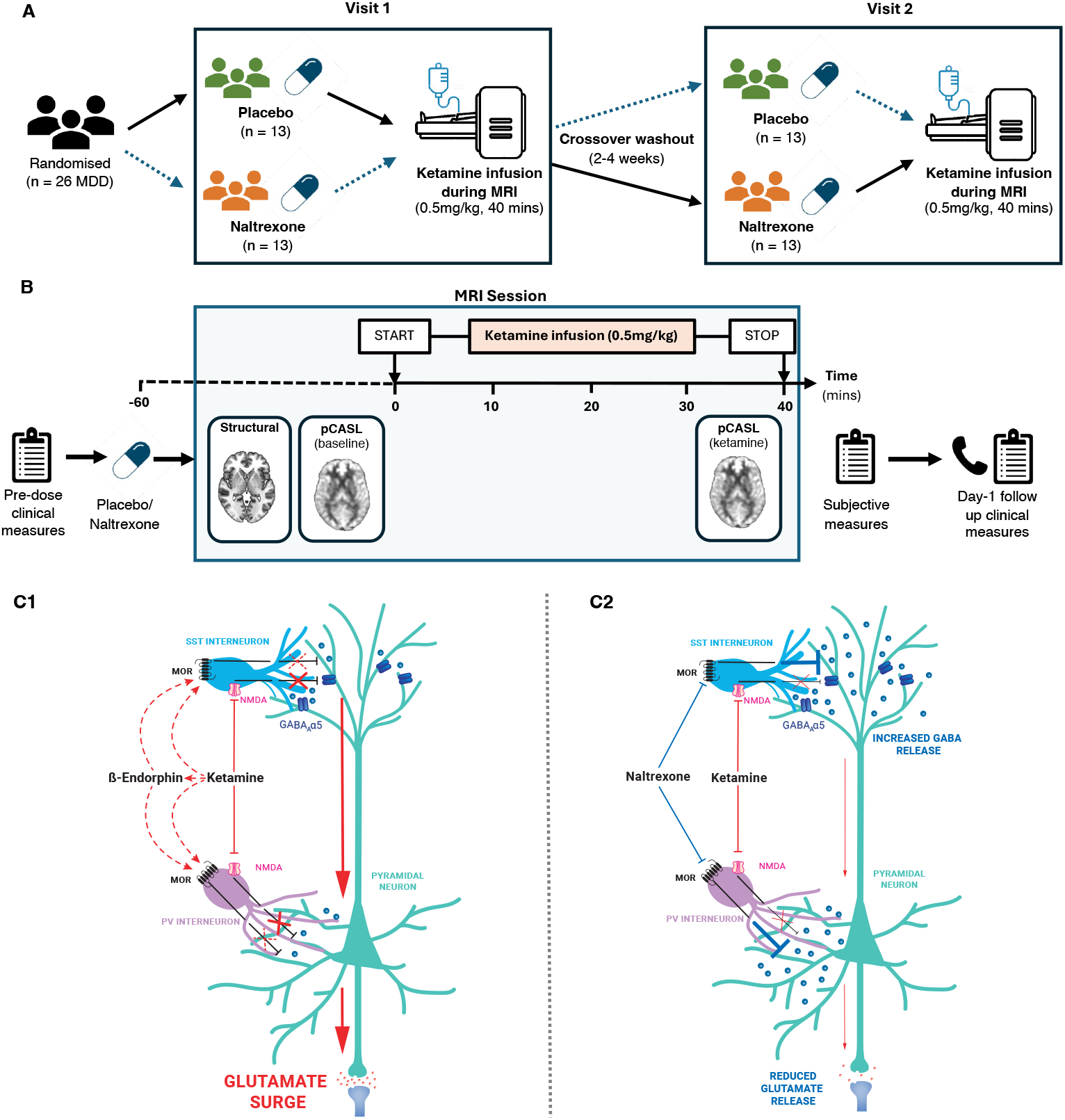
Study overview and proposed mu-opioid/GABA/glutamate model of ketamine response. (A) Twenty-six participants with MDD completed a within-subject, randomised, double-blind crossover study: oral placebo or naltrexone was given 1 h before ketamine (0.5 mg/kg over 40 min) at two visits separated by a 2–4 week washout (counterbalanced order; N=13 per sequence). (B) At each visit, clinical assessments were completed pre-dose and 24 h post-infusion. During MRI, a structural scan and baseline 3D-pCASL CBF scan were acquired, followed by ketamine infusion and a second 3D-pCASL scan during the final 10 min; subjective ratings were collected immediately post-infusion (peak effects recalled). (C) Proposed mechanism: ketamine inhibits NMDA receptors on GABAergic interneurons, disinhibiting pyramidal neurons and increasing glutamatergic activity. μ-opioid receptors (MORs) on interneurons may further contribute by suppressing inhibitory signalling. Ketamine’s potential dual role as an NMDA antagonist and a direct/indirect MOR agonist (possibly via β-endorphin release) could synergistically enhance disinhibition, leading to increased glutamate release. With naltrexone, MOR blockade may increase inhibitory tone and attenuate ketamine-related glutamatergic effects.

### Participants

Participant details have been previously reported (4). Briefly, 26 adults (18-50 years) with MDD diagnosed according to DSM-5 completed both study arms. Inclusion required a Hamilton Depression Rating Scale (HAM-D) score of at least 18 and prior inadequate response to two or more antidepressants (minimum effective dose for ≥6 weeks) or one antidepressant and a course of evidence-based psychotherapy. Regular antidepressants (≥4 weeks) were permitted, except monoamine oxidase inhibitors (MAOIs). Exclusion criteria included pregnancy, breastfeeding, schizophrenia, schizoaffective or bipolar disorder, current substance dependence, high suicide risk, prior nonresponse or intolerance to ketamine, uncontrolled medical illnesses, and MRI contraindications. Complete ASL data sets were available for 25 participants, after data from one participant was excluded due to image artifact. See

#### Supplementary

*Participant Ketamine History and Substance Use Screening* and **eTable 1** for further details and participant demographics of the included sample.

#### Imaging Methods

Detailed imaging methods are provided in the **Supplementary eMethods**. Briefly, all MRI data were acquired on a 3 Tesla General Electric (GE) Discovery MR750 scanner equipped with a 32-channel head coil. rCBF was measured at rest using 3D-pCASL with a post-labelling delay of 2025 ms and four control-label pairs collected to derive perfusion-weighted images. A proton density (PD) image was also acquired within the same series, after the control-label pairs, using identical readout parameters. The 3D Fast Spin Echo spiral multi-shot readout had eight interleaved spiral arms and 512 points per arm. Four background suppression pulses were employed. The PD image enabled computation of CBF maps in absolute, standard physiological units (mL blood/100 g tissue/min). High-resolution T1-weighted images were obtained for structural reference and spatial normalisation of the CBF maps.

### Image Processing

3D-pCASL pre-processing used the *ASAP* (Automatic Software for ASL Processing) toolbox (24) which uses Statistical Parametric Mapping (SPM12) software. PD images were co-registered to each participant’s T1-weighted image, and the resulting transformation was applied to the CBF maps. Each CBF map was then spatially normalised to MNI-152 template space, masked to exclude non-brain tissue, and spatially smoothed using a 6-mm Gaussian kernel.

### Analysis of 3D-pCASL Data

#### Analysis plan

ASL was a pre-specified secondary neuroimaging outcome in the pre-registered study which was powered a priori on the primary magnetic resonance spectroscopy outcome, reported elsewhere (4). The confirmatory ASL analyses were: (i) ketamine (infusion) vs pre-infusion baseline effects on CBF in a priori ROIs and at the whole-brain level; and (ii) the ketamine × pretreatment (naltrexone vs placebo) interaction for these outcomes, analysed with linear mixed-effects (ROI and global) and voxel-wise models with family-wise error control. With N = 25 and α = 0.05 (two-sided), our within-subject design has ~80% power to detect effects of Cohen’s d ≈ 0.58 on rCBF, in line with acute ketamine effects reported previously (15); smaller effects may go undetected. Associations with clinical/subjective measures and spatial correlations with receptor density maps were exploratory and hypothesis-generating, intended to contextualise potential mechanisms and to be interpreted with caution.

#### Global CBF

Global CBF values were calculated by extracting mean signal from a grey matter mask using *MarsBaR*. White matter was excluded because ASL has well-established low sensitivity in white-matter tissue. A linear mixed-effects (LME) model was used to assess the main effects of ketamine, pretreatment condition (naltrexone vs. placebo), and their interaction on global CBF, with participant ID included as a random effect. Analyses were implemented using R software (version 4.2.1) with the *nlme* package.

#### Region of interest (ROI)

We selected nine pre-defined regions, based on prior literature, that have been implicated in ketamine’s effects: sgACC area 25 (left and right), sgACC area 32, pgACC, dACC (left and right), thalamus, hippocampus, and anterior insula (see **Supplementary eMethods** and **eFigure 1**). Mean rCBF values for each ROI were extracted, and LME models were applied to examine the main effect of ketamine and ketamine-by-naltrexone interaction, controlling for global grey matter CBF. Multiple comparisons were corrected using the Benjamini-Hochberg method (q= 0.05) across all ROI tests reported (9 ROIs × 3 contrasts; 27 tests).

#### Whole-Brain Voxel-Wise Analyses

Whole-brain voxel-wise analyses were conducted in SPM12 using a flexible factorial design. This approach allowed for the assessment of main and interaction effects of ketamine and pretreatment condition on CBF. Age and sex were included as covariates to account for their influence on CBF. For whole-brain analyses, global normalisation (global grey-matter CBF adjustment) was performed by including each scan’s mean grey-matter CBF as a covariate in the SPM model. Results without this covariate are provided in the Supplementary materials. Statistical maps were thresholded at *p* < 0.05, voxel-wise (peak-level) FWE-corrected.

#### Associations with Clinical and Subjective Measures

We used partial Pearson correlations to test whether baseline ROI rCBF (pre-ketamine) was associated with day-1 changes in clinical scales (MADRS, QIDS-SR, Snaith-Hamilton Pleasure Scale (SHAPS), Temporal Experience of Pleasure Scale - Anticipatory (TEPS-A), and Temporal Experience of Pleasure Scale - Consummatory (TEPS-C)), adjusting for global CBF. During infusion, we computed relative rCBF (rCBF / global CBF) for each ROI and correlated these against day-1 clinical change scores, adjusting for pre-infusion relative rCBF. Analyses of acute subjective effects (CADSS; PSI subscales) followed the same approach for baseline and infusion ROI rCBF. All tests were run separately under placebo and naltrexone, with Bonferroni correction across nine ROIs (α < 0.0056). To compare placebo versus naltrexone correlations, we applied Steiger’s Z-test.

#### Receptor Density and CBF Correlation Analysis

Spatial correlations between ketamine-induced CBF changes and receptor distributions, alongside the effect of naltrexone, were examined using publicly available PET-derived receptor maps (MOR, KOR, NMDA, mGluR5, GABA_A_, and GABA_A_α5). To account for spatial autocorrelation, Spearman’s correlations between CBF contrast maps and receptor density maps were corrected using BrainSMASH. (25) (see **Supplementary eMethods** and **eFigure 2**). Given evidence for potential dopaminergic and serotonergic interactions in ketamine’s antidepressant effects (26), we also analysed dopaminergic (D_1_ and D_2_) and serotonergic PET maps (5HT_1A_, 5HT_1B_, 5HT_2A_, 5HT_4_, 5HT_6_, and 5HTT).

**Figure 2:**
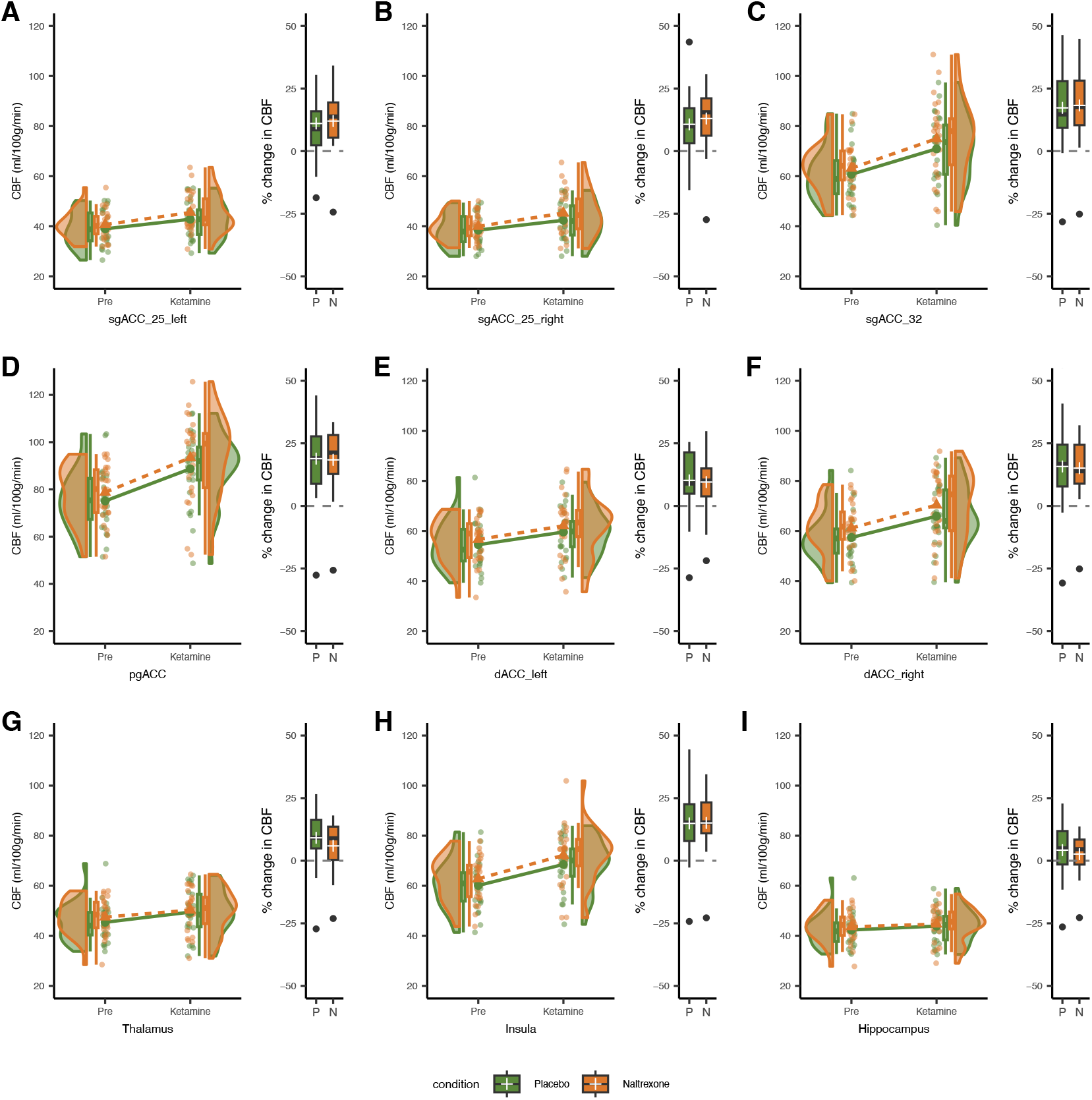
Regional CBF values and percentage change across regions of interest. CBF values are shown for the Pre (pre-infusion) and Ketamine (during infusion) scans for each pretreatment condition (placebo and naltrexone). The thick lines represent the mean values for each condition. Panels represent the following ROIs: (A) sgACC-25 (left), (B) sgACC-25 (right), (C) sgACC-32, (D) pgACC, (E) dACC (left), (F) dACC (right), (G) thalamus (bilateral), (H) insula (bilateral), and (I) hippocampus (bilateral). Boxplots to the right of each panel display the percentage change in CBF for placebo (P) and naltrexone (N) conditions. Box plot elements: Box spans the interquartile range (IQR) (25th–75th percentile), central line is the median (50th percentile), “+” marks the mean, whiskers extend to the lowest and highest values within 1.5 × IQR of the box, and values beyond that range are shown individually. Each plot shows data from participants with complete 3D-pCASL data sets (n = 25).

## Results

### Global CBF

Linear mixed-effects analysis revealed a significant main effect of ketamine on global grey-matter CBF (F_1,72_ = 18.1, p < 0.001), with increased CBF during infusion in both the placebo-plus-ketamine (mean increase = 3.75 ± 5.01 ml/100 g/min) and naltrexone-plus-ketamine (mean increase = 3.25 ± 4.25 ml/100 g/min) conditions. A significant main effect of pretreatment condition was also observed (F_1,72_ = 5.27, p = 0.025), with higher overall global CBF in the naltrexone condition (mean = 51.39 ± 8.31) compared to placebo (mean = 49.50 ± 8.88). No significant ketamine-by-pretreatment interaction was found (F_1,72_ = 0.09, p = 0.76) (**Supplementary eFigure 3**).

**Figure 3:**
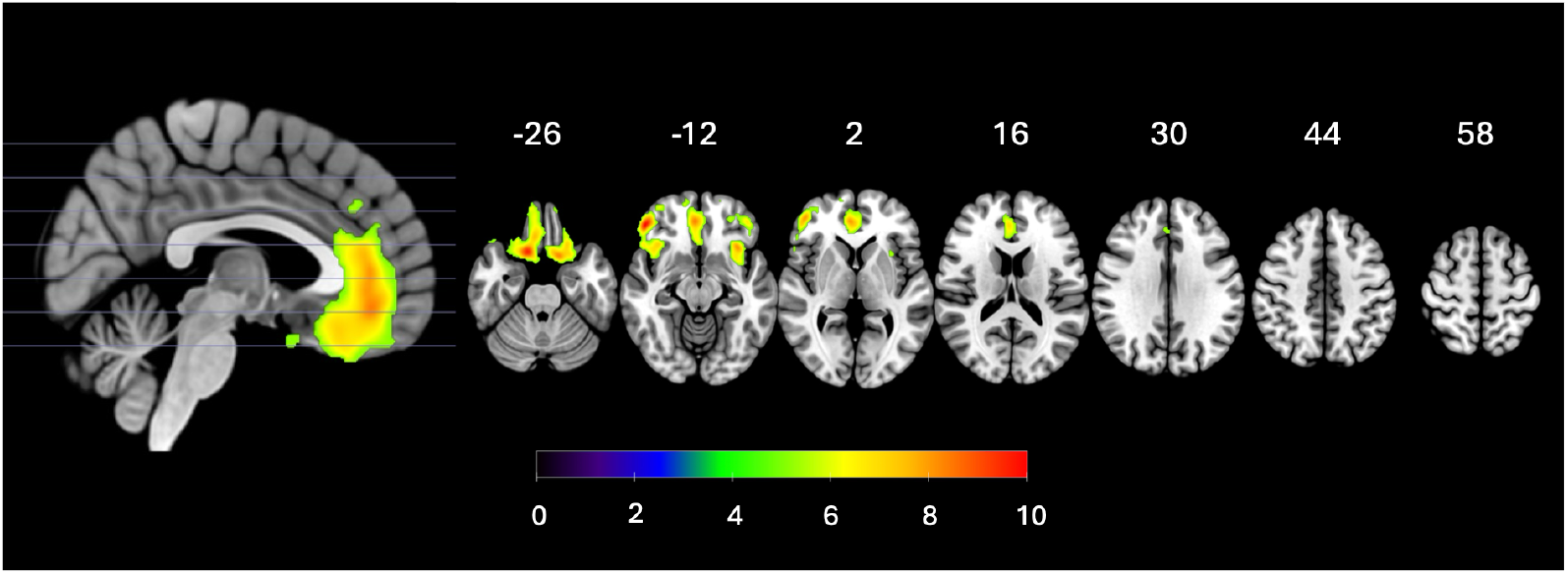
The main effect of ketamine on CBF for the whole-brain voxel-wise analysis (with global normalisation). Results overlaid on the SPM152 T1 template distributed with MRIcroGL. The MNI z-axis is shown at the top of the figure. Statistical maps were thresholded at p < 0.05, voxel-wise (peak-level) FWE-corrected. The colour bar shows the t-statistic.

### Region of interest analyses

Ketamine and pretreatment condition each had significant main effects on rCBF across all nine ROIs (FDR-adjusted p < 0.05), with increased rCBF during ketamine infusion and higher overall rCBF in the naltrexone condition. No significant interaction effects were observed (**Figure 2, Supplementary eTable 2**).

### Whole-brain analysis

Ketamine increased voxel-wise CBF in a large frontal cluster encompassing the sgACC, pgACC and dACC, anterior PFC, bilateral insula, inferior frontal gyri and orbitofrontal cortex (p < 0.05, voxel-wise FWE-corrected; **Figure 3; Supplementary eTable 3**). No significant main effects of pretreatment condition, or ketamine-by-pretreatment interactions, were observed in the whole-brain voxel-wise analysis. Results without global CBF adjustment are shown in **Supplementary eFigure 4**.

**Figure 4:**
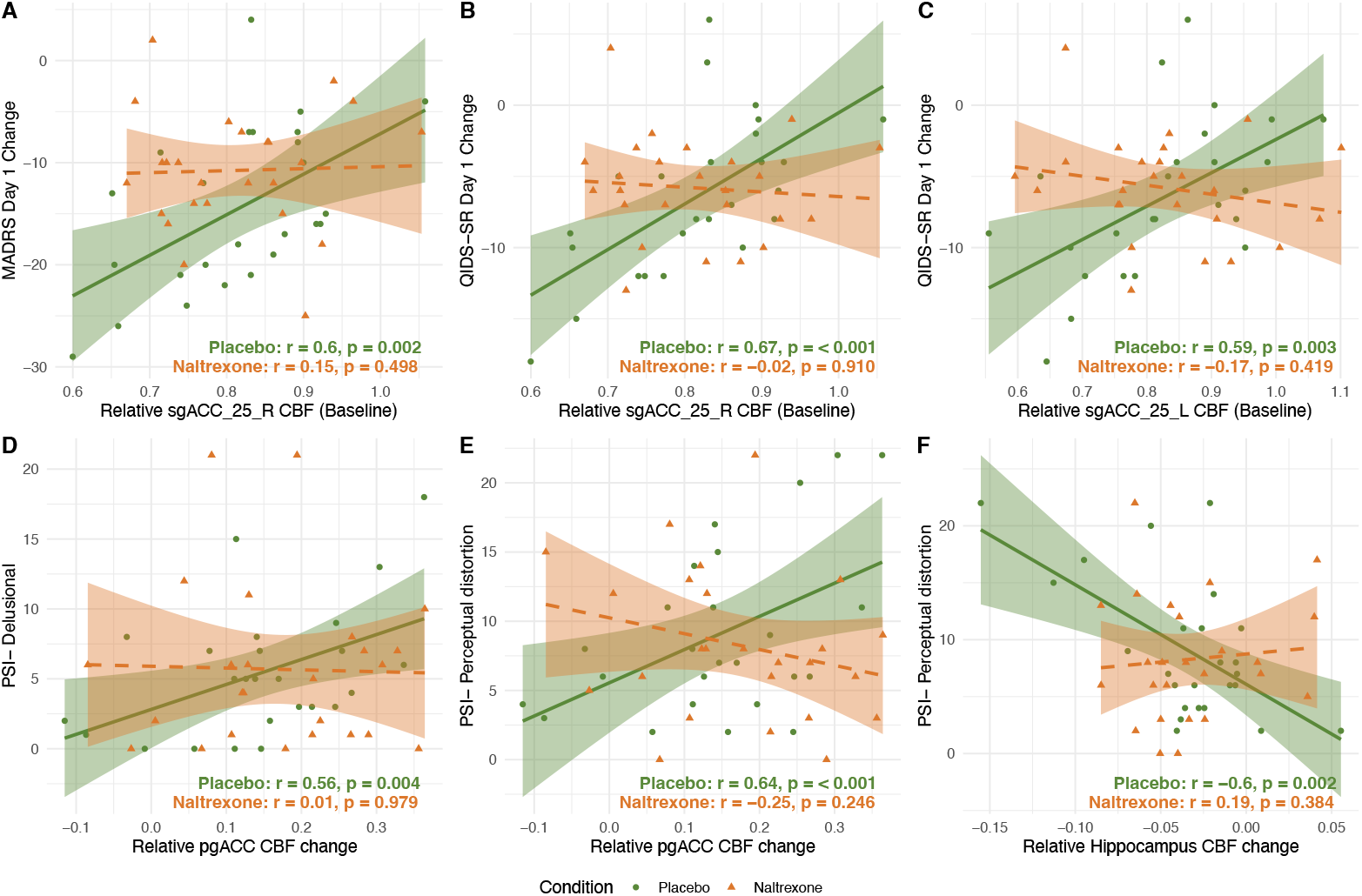
Baseline rCBF is associated with day 1 changes in clinical measures, and baseline-adjusted ketamine infusion relative rCBF is associated with subjective measure scores (placebo pretreatment condition only) (A) Baseline relative rCBF in right sgACC-25 versus day 1 MADRS change; (B) *B*aseline relative rCBF in right sgACC-25 versus day 1 QIDS-SR change; (C) Baseline relative rCBF in left sgACC-25 versus day 1 QIDS-SR change;; (D) Change in relative pgACC rCBF during ketamine infusion versus PSI delusional scores; (E) Change in relative pgACC rCBF during ketamine infusion versus PSI perceptual distortion scores; (F) Change in relative hippocampal rCBF during ketamine infusion versus PSI perceptual distortion scores. For baseline plots (A–C), r and p values are from partial correlations of ROI rCBF with outcomes, adjusting for global CBF; the x-axes in (A–C) display relative rCBF (ROI/global) for visualisation. For infusion-change plots (D–F), r and p values are from partial correlations of ketamine infusion relative ROI rCBF with subjective scores, adjusting for pre-infusion relative rCBF

### Regional CBF and clinical/subjective measure associations

In the placebo condition, significant positive partial correlations (adjusted for global CBF) were observed between baseline sgACC-25 rCBF (left and right) and day 1 improvement in depressive symptoms. (MADRS and QIDS-SR). Lower baseline sgACC-25 rCBF was associated with greater antidepressant response. These associations were absent under naltrexone (**Figure 4A–C)**. Steiger’s Z-tests demonstrated each association was significantly stronger under placebo than naltrexone: left sgACC-25–QIDS-SR (t = 2.67, p = 0.014); right sgACC-25– MADRS (t = 2.18, p = 0.040); and right sgACC-25–QIDS-SR (t = 2.55, p = 0.018).

In the placebo condition, baseline-adjusted infusion relative rCBF in the pgACC was positively correlated with PSI delusional and perceptual distortion scores, whereas baseline-adjusted infusion relative hippocampal rCBF was negatively correlated with perceptual distortion. These associations were abolished under naltrexone (**Figure 4D–F**). Steiger’s Z-tests demonstrated each association was significantly stronger under placebo than naltrexone: pgACC–PSI delusional (t = 6.12, p < 0.001), pgACC–PSI perceptual distortion (t = 8.46, p < 0.001), and hippocampus–PSI perceptual distortion (t = –5.86, p < 0.001).

A positive correlation was also found between baseline left sgACC-25 rCBF and PSI anhedonia under placebo (r = 0.625, p = 0.001), but not naltrexone (r = 0.322, p = 0.125). Conversely, baseline thalamus rCBF was negatively correlated with PSI paranoia under naltrexone (r = – 0.562, p = 0.004), but not placebo (r = –0.322, p = 0.125). However, these correlations did not differ significantly between conditions (t = 1.59, p = 0.13; t = 1.88, p = 0.073).

### ΔCBF and receptor density profile associations

Spatial correlation analyses showed a significant association between ketamine-induced ΔCBF and MOR distribution (r=0.512, pSA-corr=0.0209), and a weaker but significant correlation with mGluR5 distribution (r=0.348, pSA-corr=0.0038) (**Figure 5 and Supplementary eTable 4**). For the ketamine x naltrexone interaction contrast, significant correlations were observed with MOR, mGluR5 and GABA_A_α5 receptor distributions (r=0.539, pSA-corr=0.0194; r=0.257, pSA-corr=0.0387, and r=0.520, pSA-corr=0.0021, respectively). No significant spatial correlations were found with KOR, NMDA, or GABA_A_ receptor maps. See **Supplementary eFigure 5 and eTable 5** for dopaminergic and serotonergic receptor density associations.

**Figure 5.**
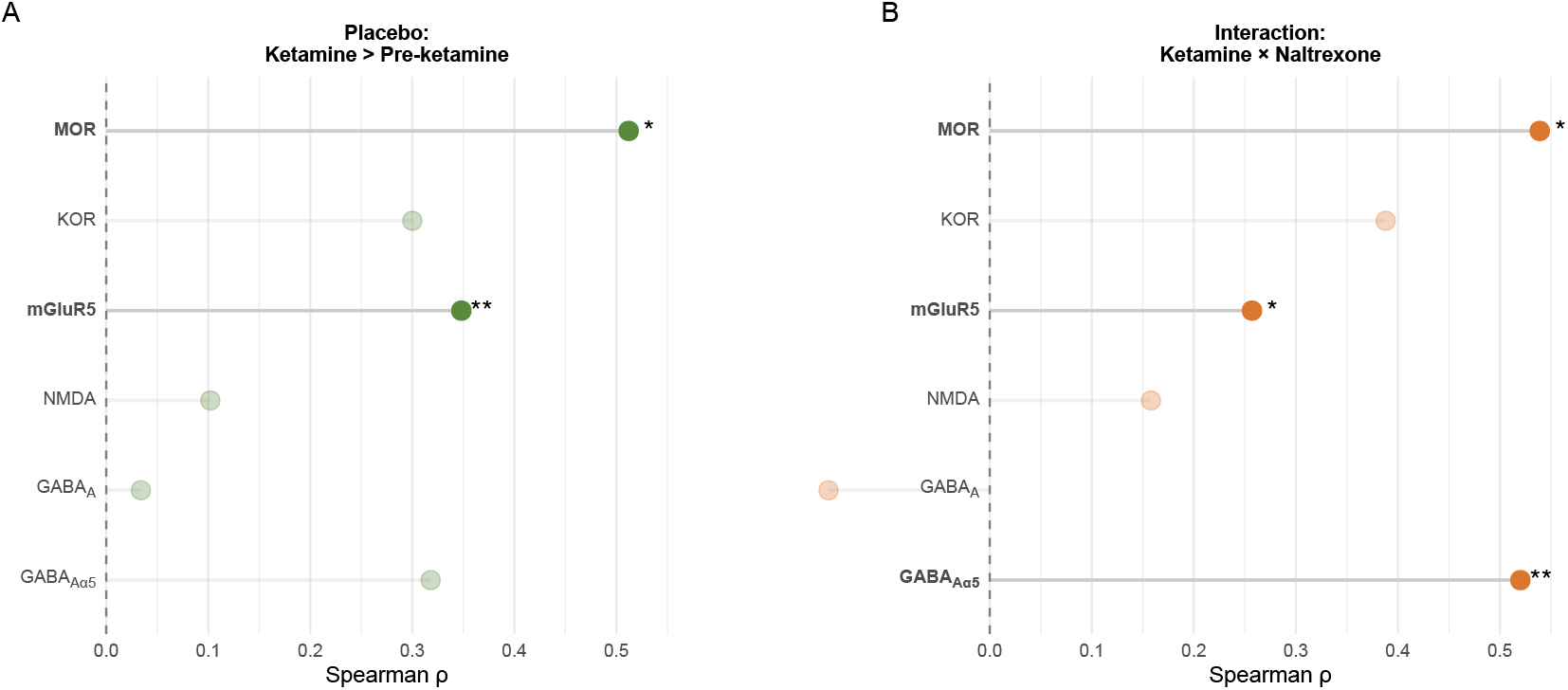
Associations between ΔCBF and receptor density profiles. (A) Main effect of ketamine under placebo (Ketamine > Pre-Ketamine) with Spearman correlations against each receptor map. (B) Ketamine × Naltrexone interaction effect on CBF with Spearman correlations against each receptor map. Asterisks denote correlations significant after spatial-autocorrelation correction (* pSA-corr < 0.05) and after additional Benjamini–Hochberg FDR correction (** FDR-corr < 0.05).

## Discussion

In this study, we used non-invasive perfusion MRI to examine the neurophysiological mechanisms underlying ketamine’s rapid antidepressant effects and explored the impact of opioid receptor antagonism using naltrexone. Consistent with our hypothesis, ketamine significantly increased rCBF in the sgACC, pgACC and dACC as well as the thalamus. Additional rCBF increases were observed in the bilateral insula, hippocampus, and frontal regions, including the anterior PFC, inferior frontal gyri, and orbitofrontal cortex. Contrary to our hypothesis, naltrexone did not significantly attenuate these ketamine-induced CBF changes in any focal region. Instead, naltrexone was associated with increased global CBF. ROI analyses indicated that baseline-adjusted infusion pgACC relative rCBF was associated with acute subjective effects, whereas baseline sgACC rCBF (adjusted for global CBF) was associated with next-day reductions in depression ratings. These associations were abolished by naltrexone pretreatment. Finally, we found significant relationships between the spatial distribution of perfusion changes induced by ketamine and opioidergic, and glutamatergic systems (MOR and mGluR5), and between the spatial distribution of the naltrexone interaction effect on ketamine induced CBF changes and opioidergic, glutamatergic and GABAergic systems (MOR, mGluR5 and GABA_A_α5).

Our findings align with previous studies in healthy volunteers, where ketamine has been shown to increase rCBF in similar regions, including anterior and subgenual areas of the cingulate, thalamus, and hippocampus (15,17,19,27,28). These regions are consistently implicated in the pathophysiology of major depression and in the therapeutic effects of antidepressant treatments (29,30). Specifically, hyperactivity in the sgACC is a well-established marker of depression, and successful treatment is often associated with normalisation of this region’s activity (30–33). Our observation that lower baseline sgACC-25 rCBF (adjusted for global CBF) was associated with greater next day antidepressant response to ketamine under placebo pretreatment is consistent with prior work suggesting that relative hyperactivity in the sgACC (Brodmann areas 24, 25, 32) is linked to poorer response to both pharmacological and psychological treatments (34–36). Together, these findings suggest that sgACC perfusion may represent a candidate biomarker of treatment outcome.

Under placebo pretreatment, infusion relative rCBF in the pgACC and hippocampus (adjusted for pre-infusion relative rCBF) was associated with acute subjective effects, including perceptual distortions and delusional thoughts. Notably, naltrexone pretreatment abolished these relationships, as well as the association between baseline sgACC-25 rCBF and delayed (24-hour) antidepressant effects. These findings suggest that the acute and delayed effects of ketamine may involve distinct neural circuits, both modulated by opioidergic systems. This differential mediation may help explain the inconsistent relationship between ketamine’s immediate subjective effects and its longer-lasting antidepressant response (37).

Naltrexone was associated with a global increase in CBF, without significant focal effects. This finding could indicate a diffuse modulatory impact of naltrexone on neurovascular function, potentially through direct effects on cerebral vasculature or interference with endorphinergic mechanisms in CBF regulation (38). A prior study using naloxone did not demonstrate similar effects (16), although methodological differences (e.g. partial brain coverage) or participant characteristics (healthy volunteers and patients with depression) may explain this discrepancy.

Our exploratory analyses revealed that the spatial distribution of ketamine-induced CBF changes was significantly correlated with receptor density maps for MOR and mGluR5. The effect of naltrexone on the spatial distribution of ketamine-induced CBF changes was associated with MOR and GABAAα5, with a more modest association with mGluR5. In vivo target specificity and state sensitivity of [_18F_]GE-179 for NMDA receptors in humans remain incompletely established (39,40), therefore the lack of correlation between ketamine’s effects on CBF and the NMDA receptor map may reflect limited or state-dependent specificity of this ligand for NMDA receptor density. The association between ketamine’s CBF effects and mGluR5 is consistent with prior evidence showing that ketamine can reduce mGluR5 availability in MDD, potentially reflecting downstream glutamatergic modulation (41). The interaction-level associations with MOR, GABAAα5 and mGluR5 suggest that mu-opioid– GABA-glutamate interplay may shape ketamine’s perfusion effects under opioid antagonism. GABA_A_α5 subunit receptors, located in cortical and limbic brain regions associated with depression, facilitate inhibition via SST-expressing interneurons on dendrite-targeting projections (42). GABAergic interneurons have been identified as the cellular trigger for ketamine’s rapid antidepressant actions, with evidence that GluN2B subunit deletion in SST interneurons blocks ketamine’s behavioural efficacy in animal models (43). MORs are also located on GABAergic interneurons (44). It has been shown that MOR expression, specifically on SST interneurons is necessary for the acute and antidepressant-like responses to the atypical antidepressant tianeptine (45). Through this circuit, activating MORs on SST interneurons can decrease their activity, providing an additional mechanism that could contribute to the disinhibition of glutamatergic pyramidal neurons.

Taken together, our findings support a speculative model whereby ketamine acts to inhibit SST interneurons, both via NMDA antagonism and via direct or indirect effects on MORs, also located on SST interneurons. This would lead to the disinhibition of pyramidal cells and an acute glutamate burst (4). In the presence of naltrexone, direct MOR antagonist effects on the SST interneurons would act to increase GABA release onto pyramidal cells dendrites, thereby increasing the inhibitory balance and attenuating ketamine-induced increases in glutamatergic activity (**Figure 1C**). This is a selective model and the full range of circuits and neurotransmitter systems at play is much more elaborate and multifaceted. Indeed, our supplementary analysis also uncovered relationships between a serotonergic receptor density map and the spatial distribution of ketamine CBF effects (5-HT1_b_) and ketamine – naltrexone interaction effects with dopaminergic and serotonergic maps (D_1_, D_2_, 5-HT_1a_, 5-HT_4_ and 5-HTT) (**Supplementary eFigure 5 and eTable 5**). This further supports the view that opioid modulation is not an isolated effect but involves interactions with other neurotransmitter systems, including glutamatergic, GABAergic, dopaminergic, and serotonergic, all implicated in ketamine’s antidepressant actions (26).

### Strengths and Limitations

This study is the first to explore the impact of opioid receptor antagonism on acute rCBF changes induced by ketamine in patients with MDD using a randomised, double-blind, crossover design. The use of 3D-pCASL imaging provided quantitative, non-invasive measurements of rCBF, enhancing the reliability of our findings. Our exploratory analysis linking CBF changes with receptor density profiles offers novel insights into the potential neurotransmitter systems involved in ketamine’s effects.

Several limitations should be acknowledged. ASL was a secondary outcome, and the study was powered for the primary magnetic resonance spectroscopy measure rather than for detecting main or interaction rCBF effects. Although our within-subject design had good sensitivity for moderate ketamine effects, it was likely underpowered to detect smaller rCBF changes or more modest ketamine–naltrexone interactions, so null findings should be interpreted with caution. Because our pCASL protocol used a single post-labelling delay, we could not estimate arterial transit time (ATT); ketamine-related ATT shifts could therefore bias CBF quantification. We used a relatively long post-labelling delay (2025 ms) to reduce ATT sensitivity, but residual confounding remains possible. Without a healthy control group, we are limited to determine whether the observed findings are specific to individuals with MDD. The lack of a placebo infusion arm also prevents us from conclusively attributing observed rCBF changes to ketamine rather than non-specific effects. While age and sex were included as covariates, other factors such as menstrual cycle phase in female participants, BMI and concurrent medications were not systematically controlled in our within-subject analyses. In addition, although heart rate, SpO_2_, and respiratory rate were monitored continuously for safety, these values were not recorded, so we could not formally test or adjust for physiological confounds, and subtle cardio-respiratory contributions to rCBF changes cannot be excluded. Finally, receptor associations were based on group level PET maps from healthy populations, which may not reflect distributions in MDD (e.g., reduced MOR and mGluR5 density in MDD (46,47)). Future studies using individualised receptor mapping may provide greater precision.

## Conclusion

This study demonstrates that ketamine administration in MDD patients increases rCBF in regions critical for emotional processing, including the sgACC, pgACC, dACC, thalamus, insula, and hippocampus. Baseline rCBF was associated with antidepressant response, whereas baseline-adjusted infusion relative rCBF was associated with acute subjective effects. However, these associations were disrupted by naltrexone pretreatment. Together, our findings suggest that ketamine’s impact on brain perfusion is not solely mediated by glutamatergic mechanisms but is also influenced by interactions with opioidergic and other neurotransmitter systems. Consistent with this, spatial correlations between ketamine-induced CBF changes and receptor density maps underscore the complexity of ketamine’s neurobiology, supporting a dynamic interplay among multiple systems, including, but not limited to glutamatergic, opioidergic, and GABAergic pathways.

## Data Availability

Data produced in the present study are available upon reasonable request to the authors.

## Acknowledgements

We thank all study participants, and the research nurses, radiographers, and support staff at the NIHR King’s Clinical Research Facility for their invaluable assistance. This is independent research funded by a Medical Research Council Clinical Research Training Fellowship (grant number MR/T028084/1 awarded to L.A.J) and carried out at the NIHR (National Institute for Health and Care Research) Maudsley Biomedical Research Centre (BRC). The views expressed are those of the authors and not necessarily those of the Medical Research Council, the National Institute for Health and Care Research or the Department of Health and Social Care.

## Competing Interests

**L.A.J:** The author declared no potential conflicts of interest with respect to the research, authorship, and/or publication of this article. **O.O:** The author declared no potential conflicts of interest with respect to the research, authorship, and/or publication of this article. **F.O.Z:** The author declared no potential conflicts of interest with respect to the research, authorship, and/or publication of this article. **J.M.S:** The author declared no potential conflicts of interest with respect to the research, authorship, and/or publication of this article. **A.H.Y:** Employed by Imperial College London; Honorary Consultant South London and Maudsley NHS Foundation Trust (NHS UK). Editor of Journal of Psychopharmacology and Deputy Editor, BJPsych Open. Paid lectures and advisory boards for the following companies with drugs used in affective and related disorders: Flow Neuroscience, Novartis, Roche, Janssen, Takeda, Noema pharma, Compass, Astrazenaca, Boehringer Ingelheim, Eli Lilly, LivaNova, Lundbeck, Sunovion, Servier, Livanova, Janssen, Allegan, Bionomics, Sumitomo Dainippon Pharma, Sage, Neurocentrx. Principal Investigator in the Restore-Life VNS registry study funded by LivaNova. Principal Investigator on ESKETINTRD3004: “An Open-label, Long-term, Safety and Efficacy Study of Intranasal Esketamine in Treatment-resistant Depression.” Principal Investigator on “The Effects of Psilocybin on Cognitive Function in Healthy Participants”. Principal Investigator on “The Safety and Efficacy of Psilocybin in Participants with Treatment-Resistant Depression (P-TRD)”. Principal Investigator on ‘‘A Double-Blind, Randomized, Parallel-Group Study with Quetiapine Extended Release as Comparator to Evaluate the Efficacy and Safety of Seltorexant 20 mg as Adjunctive Therapy to Antidepressants in Adult and Elderly Patients with Major Depressive Disorder with Insomnia Symptoms Who Have Responded Inadequately to Antidepressant Therapy.’’ (Janssen). Principal Investigator on ‘‘An Open-label, Long-term, Safety and Efficacy Study of Aticaprant as Adjunctive Therapy in Adult and Elderly Participants with Major Depressive Disorder (MDD).’’ (Janssen). Principal Investigator on ‘‘A Randomized, Double-blind, Multicentre, Parallel-group, Placebo-controlled Study to Evaluate the Efficacy, Safety, and Tolerability of Aticaprant 10 mg as Adjunctive Therapy in Adult Participants with Major Depressive Disorder (MDD) with Moderate-to-severe Anhedonia and Inadequate Response to Current Antidepressant Therapy.’’ Principal Investigator on ‘‘A Study of Disease Characteristics and Real-life Standard of Care Effectiveness in Patients with Major Depressive Disorder (MDD) With Anhedonia and Inadequate Response to Current Antidepressant Therapy Including an SSRI or SNR.’’ (Janssen). UK Chief Investigator for Compass; COMP006 & COMP007 studies. UK Chief Investigator for Novartis MDD study MIJ821A12201. Grant funding (past and present): NIMH (USA); CIHR (Canada); NARSAD (USA); Stanley Medical Research Institute (USA); MRC (UK); Wellcome Trust (UK); Royal College of Physicians (Edin); BMA (UK); UBC-VGH Foundation (Canada); WEDC (Canada); CCS Depression Research Fund (Canada); MSFHR (Canada); NIHR (UK). Janssen (UK) EU Horizon 2020. No shareholdings in pharmaceutical companies. **M.A.M:** Has current funding from Nxera, Takeda and Lundbeck and has acted as an advisor for Boehringer Ingelheim, Neurocentrx, Quolet Pharmaceuticals and Nxera.

## Notes

### Clinical Trial

NCT04977674

### Author Declarations

Ethical approval was obtained from the London - City & East Research Ethics Committee (Reference: 21/LO/0334) and all participants provided written informed consent. This is a mechanistic, randomised, double-blind, placebo-controlled crossover study and not classified as a Clinical Trial of an Investigational Medicinal Product in the UK. The study received ethical approval as a basic science study in humans under EU Directive 2001/20/EC. However, the study was still registered at ClinicalTrials.gov (NCT04977674).

### Summary of Updates

Version updated to correct receptor mapping analysis results after template mismatch. This includes updates to Figure 5 and supplementary materials.

## References

1. Jelen LA, Stone JM. Ketamine for depression. Int Rev Psychiatry. 2021/02/12 edn. 2021 Feb 11;1–32. Located at: 33569971. doi:10.1080/09540261.2020.1854194

2. Zavaliangos-Petropulu A, Al-Sharif NB, Taraku B, Leaver AM, Sahib AK, Espinoza RT, et al. Neuroimaging-Derived Biomarkers of the Antidepressant Effects of Ketamine. Biological Psychiatry: Cognitive Neuroscience and Neuroimaging. 2023 Apr 1;8(4):361–86. doi:10.1016/j.bpsc.2022.11.005

3. Zanos P, Moaddel R, Morris PJ, Riggs LM, Highland JN, Georgiou P, et al. Ketamine and Ketamine Metabolite Pharmacology: Insights into Therapeutic Mechanisms. Pharmacol Rev. 2018/06/28 edn. 2018 Jul;70(3):621–60. Located at: 29945898. doi:10.1124/pr.117.015198

4. Jelen LA, Lythgoe DJ, Stone JM, Young AH, Mehta MA. Effect of naltrexone pretreatment on ketamine-induced glutamatergic activity and symptoms of depression: a randomized crossover study. Nat Med. 2025 Jul 24;1–9. doi:10.1038/s41591-025-03800-w

5. Klein ME, Chandra J, Sheriff S, Malinow R. Opioid system is necessary but not sufficient for antidepressive actions of ketamine in rodents. Proc Natl Acad Sci US A. 2020/01/17 edn. 2020 Feb 4;117(5):2656–62. Located at: 31941713. doi:10.1073/pnas.1916570117

6. Williams NR, Heifets BD, Blasey C, Sudheimer K, Pannu J, Pankow H, et al. Attenuation of Antidepressant Effects of Ketamine by Opioid Receptor Antagonism. Am J Psychiatry. 2018/08/30 edn. 2018 Dec 1;175(12):1205–15. Located at: 30153752. doi:10.1176/appi.ajp.2018.18020138

7. Di Ianni T, Ewbank SN, Levinstein MR, Azadian MM, Budinich RC, Michaelides M, et al. Sex dependence of opioid-mediated responses to subanesthetic ketamine in rats. Nat Commun. 2024 Jan 30;15(1):893. doi:10.1038/s41467-024-45157-7

8. Jiang C, DiLeone RJ, Pittenger C, Duman RS. The endogenous opioid system in the medial prefrontal cortex mediates ketamine’s antidepressant-like actions. Transl Psychiatry. 2024 Feb 12;14(1):90. doi:10.1038/s41398-024-02796-0 PubMed PMID: 38346984; PubMed Central PMCID: PMC10861497.

9. Zhang F, Hillhouse TM, Anderson PM, Koppenhaver PO, Kegen TN, Manicka SG, et al. Opioid receptor system contributes to the acute and sustained antidepressant-like effects, but not the hyperactivity motor effects of ketamine in mice. Pharmacol Biochem Behav. 2021/07/06 edn. 2021 Jul 2;173228. Located at: 34224734. doi:10.1016/j.pbb.2021.173228

10. Pomrenze MB, Vaillancourt S, Llorach P, Rijsketic DR, Casey AB, Gregory N, et al. Ketamine evokes acute behavioral effects via μ-opioid receptor expressing neurons of the central amygdala. Biological Psychiatry. 2025 May 5. doi:10.1016/j.biopsych.2025.04.020

11. Yoon G, Petrakis IL, Krystal JH. Association of Combined Naltrexone and Ketamine With Depressive Symptoms in a Case series of Patients With Depression and Alcohol Use Disorder. JAMA Psychiatry. 2019 Mar 1;76(3):337. doi:10.1001/jamapsychiatry.2018.3990

12. Zhang K, Hashimoto K. Lack of Opioid System in the Antidepressant Actions of Ketamine. Biol Psychiatry. 2018/12/14 edn. 2019 Mar 15;85(6):e25–7. Located at: 30545521. doi:10.1016/j.biopsych.2018.11.006

13. Attwell D, Buchan AM, Charpak S, Lauritzen M, MacVicar BA, Newman EA. Glial and neuronal control of brain blood flow. Nature. 2010 Nov;468(7321):7321. doi:10.1038/nature09613

14. Petcharunpaisan S, Ramalho J, Castillo M. Arterial spin labeling in neuroimaging. World J Radiol. 2010/12/17 edn. 2010 Oct 28;2(10):384–98. Located at: 21161024. doi:10.4329/wjr.v2.i10.384

15. Pollak TA, De Simoni S, Barimani B, Zelaya FO, Stone JM, Mehta MA. Phenomenologically distinct psychotomimetic effects of ketamine are associated with cerebral blood flow changes in functionally relevant cerebral foci: a continuous arterial spin labelling study. Psychopharmacology (Berl). 2015/10/07 edn. 2015 Dec;232(24):4515–24. Located at: 26438425. doi:10.1007/s00213-015-4078-8

16. Zelaya FO, Zois E, Muller-Pollard C, Lythgoe DJ, Lee S, Andrews C, et al. The response to rapid infusion of fentanyl in the human brain measured using pulsed arterial spin labelling. MAGMA. 2011/11/25 edn. 2012 Apr;25(2):163–75. Located at: 22113518. doi:10.1007/s10334-011-0293-4

17. Bojesen KB, Andersen KA, Rasmussen SN, Baandrup L, Madsen LM, Glenthoj BY, et al. Glutamate Levels and Resting Cerebral Blood Flow in Anterior Cingulate Cortex Are Associated at Rest and Immediately Following Infusion of S-Ketamine in Healthy Volunteers. Front Psychiatry. 2018/02/23 edn. 2018;9:22. Located at: 29467681. doi:10.3389/fpsyt.2018.00022

18. Holcomb HH, Lahti AC, Medoff DR, Cullen T, Tamminga CA. Effects of Noncompetitive NMDA Receptor Blockade on Anterior Cingulate Cerebral Blood Flow in Volunteers with Schizophrenia. Neuropsychopharmacol. 2005 Dec; 30(12):12. doi:10.1038/sj.npp.1300824

19. Långsjö JW, Kaisti KK, Aalto S, Hinkka S, Aantaa R, Oikonen V, et al. Effects of Subanesthetic Doses of Ketamine on Regional Cerebral Blood Flow, Oxygen Consumption, and Blood Volume in Humans. Anesthesiology. 2003 Sep 1;99(3):614–23. doi:10.1097/00000542-200309000-00016

20. Rowland LM, Beason-Held L, Tamminga CA, Holcomb HH. The interactive effects of ketamine and nicotine on human cerebral blood flow. Psychopharmacology. 2010 Mar 1;208(4):575–84. doi:10.1007/s00213-009-1758-2

21. Gärtner M, Weigand A, Meiering MS, Weigner D, Carstens L, Keicher C, et al. Region- and time-specific effects of ketamine on cerebral blood flow: a randomized controlled trial. Neuropsychopharmacol. 2023 May 25;1–7. doi:10.1038/s41386-023-01605-4

22. Gonzalez S, Vasavada MM, Njau S, Sahib AK, Espinoza R, Narr KL, et al. Acute changes in cerebral blood flow after single-infusion ketamine in major depression: A pilot study. Neurology, Psychiatry and Brain Research. 2020 Dec 1:38:5–11. doi:10.1016/j.npbr.2020.08.006

23. Sahib AK, Loureiro JR, Vasavada M, Kubicki A, Joshi S, Wang K, et al. Single and repeated ketamine treatment induces perfusion changes in sensory and limbic networks in major depressive disorder. Eur Neuropsychopharmacol. 2020 Apr;33:89–100. doi:10.1016/j.euroneuro.2020.01.017 PubMed PMID: 32061453; PubMed Central PMCID: PMC8869841.

24. Mato Abad V, García-Polo P, O’Daly O, Hernández-Tamames JA, Zelaya F. ASAP (Automatic Software for ASL Processing): A toolbox for processing Arterial Spin Labeling images. Magnetic Resonance Imaging. 2016 Apr 1;34(3):334–44. doi:10.1016/j.mri.2015.11.002

25. Burt JB, Helmer M, Shinn M, Anticevic A, Murray JD. Generative modeling of brain maps with spatial autocorrelation. Neurolmage. 2020 Oct 15;220:117038. doi:10.1016/j.neuroimage.2020.117038

26. Jelen LA, Young AH, Stone JM. Ketamine: A tale of two enantiomers. J Psychopharmacol. 2020/11/07 edn. 2021 Feb;35(2):109–23. doi:10.1177/0269881120959644 Located at: 33155503.

27. Bryant JE, Frölich M, Tran S, Reid MA, Lahti AC, Kraguljac NV. Ketamine induced changes in regional cerebral blood flow, interregional connectivity patterns, and glutamate metabolism. Journal of Psychiatric Research. 2019 Oct 1;117:108–15. doi:10.1016/j.jpsychires.2019.07.008

28. Holcomb HH, Lahti AC, Medoff DR, Weiler M, Tamminga CA. Sequential Regional Cerebral Blood Flow Brain Scans Using PET with H2150 Demonstrate Ketamine Actions in CNS Dynamically. Neuropsychopharmacol. 2001 Aug;25(2):2. doi:10.1016/S0893-133X(01)00229-9

29. Drevets WC, Savitz J, Trimble M. The subgenual anterior cingulate cortex in mood disorders. CNS Spectr. 2008/08/16 edn. 2008 Aug;13(8):663–81. Located at: 18704022

30. Mayberg HS, Lozano AM, Voon V, McNeely HE, Seminowicz D, Hamani C, et al. Deep brain stimulation for treatment-resistant depression. Neuron. 2005/03/08 edn. 2005 Mar 3;45(5):651–60. Located at: 15748841. doi:10.1016/j.neuron.2005.02.014

31. Kennedy SH, Konarski JZ, Segal ZV, Lau MA, Bieling PJ, Mcintyre RS, et al. Differences in brain glucose metabolism between responders to CBT and venlafaxine in a 16-week randomized controlled trial. Am J Psychiatry. 2007 May; 164(5):778–88. doi:10.1176/ajp.2007.164.5.778 PubMed PMID: 17475737.

32. Mayberg HS. Targeted electrode-based modulation of neural circuits for depression. J Clin Invest. 2009 Apr;119(4):717–25. doi:10.1172/JCI38454 PubMed PMID: 19339763; PubMed Central PMCID: PMC2662569.

33. Mayberg HS, Brannan SK, Tekell JL, Silva JA, Mahurin RK, McGinnis S, et al. Regional metabolic effects of fluoxetine in major depression: serial changes and relationship to clinical response. Biological Psychiatry. 2000 Oct 15;48(8):830–43. doi:10.1016/S0006-3223(00)01036-2

34. Brody AL, Saxena S, Stoessel P, Gillies LA, Fairbanks LA, Alborzian S, et al. Regional brain metabolic changes in patients with major depression treated with either paroxetine or interpersonal therapy: preliminary findings. Arch Gen Psychiatry. 2001 Jul;58(7):631–40. doi:10.1001/archpsyc.58.7.631 PubMed PMID: 11448368.

35. McGrath CL, Kelley ME, Dunlop BW, Holtzheimer PE, Craighead WE, Mayberg HS. Pretreatment brain states identify likely nonresponse to standard treatments for depression. Biol Psychiatry. 2014 Oct 1;76(7):527–35. doi:10.1016/j.biopsych.2013.12.005 PubMed PMID: 24462230; PubMed Central PMCID: PMC4063885.

36. Konarski JZ, Kennedy SH, Segal ZV, Lau MA, Bieling PJ, McIntyre RS, et al. Predictors of nonresponse to cognitive behavioural therapy or venlafaxine using glucose metabolism in major depressive disorder. J Psychiatry Neurosci. 2009/05/19 edn. 2009 May;34(3):175–80. Located at: 19448846

37. Ballard ED, Zarate CA. The role of dissociation in ketamine’s antidepressant effects. Nat Commun. 2020 Dec 22;11(1):1. doi:10.1038/s41467-020-20190-4

38. Benyó Z, Wahl M. Opiate receptor-mediated mechanisms in the regulation of cerebral blood flow. Cerebrovasc Brain Metab Rev. 1996;8(4):326–57. PubMed PMID: 8969868.

39. McGinnity CJ, Hammers A, Riaño Barros DA, Luthra SK, Jones PA, Trigg W, et al. Initial evaluation of 18F-GE-179, a putative PET Tracer for activated N-methyl D-aspartate receptors. J Nucl Med. 2014 Mar;55(3):423–30. doi:10.2967/jnumed.113.130641 PubMed PMID: 24525206.

40. Galovic M, Erlandsson K, Fryer TD, Hong YT, Manavaki R, Sari H, et al. Validation of a combined image derived input function and venous sampling approach for the quantification of [18F]GE-179 PET binding in the brain. NeuroImage. 2021 Aug 15:237:118194. doi:10.1016/j.neuroimage.2021.118194

41. Esterlis I, DellaGioia N, Pietrzak RH, Matuskey D, Nabulsi N, Abdallah CG, et al. Ketamine-induced reduction in mGluR5 availability is associated with an antidepressant response: an [C] ABP688 and PET imaging study in depression. Mol Psychiatry. 2018 Apr 1;23(4):824–32. doi:10.1038/mp.2017.58 PubMed PMID: 28397841; PubMed Central PMCID: PMC5636649.

42. Fee C, Banasr M, Sibille E. Somatostatin-Positive Gamma-Aminobutyric Acid Interneuron Deficits in Depression: Cortical Microcircuit and Therapeutic Perspectives. Biological Psychiatry. 2017 Oct;82(8):549–59. doi:10.1016/j.biopsych.2017.05.024

43. Gerhard DM, Pothula S, Liu RJ, Wu M, Li XY, Girgenti MJ, et al. GABA interneurons are the cellular trigger for ketamine’s rapid antidepressant actions. J Clin Invest. 2019/11/20 edn. 2020 Mar 2;130(3):1336–49. Located at: 31743111. doi:10.1172/JCI130808

44. Jiang C, Wang X, Le Q, Liu P, Liu C, Wang Z, et al. Morphine coordinates SST and PV interneurons in the prelimbic cortex to disinhibit pyramidal neurons and enhance reward. Mol Psychiatry. 2021 Apr;26(4):4. doi:10.1038/s41380-019-0480-7

45. Han J, Andreu V, Langreck C, Pekarskaya EA, Grinnell SG, Allain F, et al. Mu opioid receptors on hippocampal GABAergic interneurons are critical for the antidepressant effects of tianeptine. Neuropsychopharmacology. 2022 Jun;47(7):1387–97. doi:10.1038/s41386-021-01192-2 PubMed PMID: 34593976; PubMed Central PMCID: PMC9117297.

46. Deschwanden A, Karolewicz B, Feyissa AM, Treyer V, Ametamey SM, Johayem A, et al. Reduced metabotropic glutamate receptor 5 density in major depression determined by [(11)C)ABP688 PET and postmortem study. Am J Psychiatry. 2011 Jul;168(7):727–34. doi:10.1176/appi.ajp.2011.09111607 PubMed PMID: 21498461; PubMed Central PMCID: PMC3129412.

47. Kennedy SE, Koeppe RA, Young EA, Zubieta JK. Dysregulation of endogenous opioid emotion regulation circuitry in major depression in women. Arch Gen Psychiatry. 2006/11/08 edn. 2006 Nov;63(11):1199–208. Located at: 17088500. doi:10.1001/archpsyc.63.11.1199

