## Supplementary Material for "Regional blood flow signatures of opioidergic modulation of ketamine in major depressive disorder: a randomised crossover study"

### eMethods

**eTable 1: Participant demographics for sample with complete ASL data sets included in analyses (n = 25).**

| Characteristic | | Sample (n = 25) |
|---|---|---|
| **Age – yr (mean (SD))** | — | 35.08 (7.65) |
| **Sex – no. (%)** | Male | 13 (52.0%) |
| | Female | 12 (48.0%) |
| **Race/Ethnicity – no. (%)** | Asian/Asian British – Indian | 2 (8.0%) |
| | Asian/Asian British – Other | 2 (8.0%) |
| | Black/Black British – African | 2 (8.0%) |
| | Black/Black British – Caribbean | 1 (4.0%) |
| | White Irish | 2 (8.0%) |
| | White Other | 4 (16.0%) |
| | White UK | 12 (48.0%) |
| **Employed – no. (%)** | No | 6 (24.0%) |
| | Student | 3 (12.0%) |
| | Yes | 16 (64.0%) |
| **BMI kg/m$^2$ (mean (SD))** | — | 24.02 (2.63) |
| **Age at first MDD onset – yr (mean (SD))** | — | 22.28 (8.23) |
| **Duration of episode – yr (median [IQR])** | — | 3.50 [3.00, 12.00] |
| **Recurrent MDD – no. (%)** | No | 14 (56.0%) |
| | Yes | 11 (44.0%) |
| **Number of antidepressant failures – no. (%)** | 1–2 | 15 (60.0%) |
| | 3–4 | 6 (24.0%) |
| | 5–6 | 3 (12.0%) |
| | 7–10 | 1 (4.0%) |
| **Psychological therapy trialled – no. (%)** | Yes | 25 (100.0%) |
| **Current antidepressant treatment – no. (%)** | Yes | 11 (44.0%) |
| | No | 14 (56.0%) |
| **HAM-D at screening (mean (SD))** | — | 21.72 (2.76) |

**Participant Ketamine History and Substance Use Screening**

One participant had received a clinically administered course of IV ketamine ~3 years before enrolment, all other participants were ketamine naive. No recreational ketamine use was reported. Participants were screened to exclude current drug/alcohol dependence and any positive urine drug screen (including ketamine, opiates, methadone, cocaine, amphetamines, benzodiazepines, cannabinoids) or positive breath alcohol at screening or dosing visits. Participants were required to abstain from alcohol from 24 hours before until 24 hours after each dosing and refrain from caffeine and nicotine on imaging visit days.

**Image acquisition**

The data were collected at 3 Tesla on a General Electric (GE) Discovery MR750 magnetic resonance scanner equipped with a 32-channel head coil. A high-resolution sagittal T1-weighted (T1-w) 3D sagittal inversion recovery prepared spoiled gradient echo (IR-SPGR) scan was initially acquired (TR = 7.35 ms, TE = 3.04 ms, TI = 400 ms, FOV = 270 mm, flip-angle = 11°, matrix size = 256 × 256, slice thickness = 1.2 mm, 196 slices). For the measurement of CBF, ASL data were acquired using a 3D pseudo-continuous Arterial Spin Labelling (3D-pCASL) sequence. Four control-label pairs were acquired to derive a perfusion-weighted difference image. Arterial blood was labelled by applying a 1525 ms series of Hanning shaped radiofrequency pulses (500 µs duration per pulse, peak-to-peak gap 1500 µs) in the presence of a net magnetic field gradient along the flow direction (the z-axis of the magnet). After a post-labelling delay of 2025 ms, a whole brain volume was read using a 3D interleaved 'stack-of-spirals' Fast Spin Echo readout that consisted of eight interleaved spiral arms in the in-plane direction with 512 points per spiral interleave. The acquired images had 54 axial slice locations of 3 mm thickness and an in-plane FOV of 240 x 240 mm after transformation to a rectangular matrix (TE = 11.088, TR = 5115 ms, flip angle = 111°). Four background suppression pulses were used to minimise static tissue signal at the time of image acquisition. A proton density image, using the same acquisition parameters, was acquired at the end of the sequence to allow computation of quantitative individual CBF maps in conventional physiological units (mL blood/100 g tissue/min) from the perfusion-weighted image derived from the averaged difference image from the control-label pairs. The total acquisition time of the 3D-pCASL sequence was 6 min and 20 s.

Voxel wise computation of CBF was performed by the scanner software, using the formula recommended by the ASL consensus article (Alsop et al., 2015):

$$CBF = 6000 \frac{e^{w/T_{1a}}}{2\varepsilon T_{1a}(1 - e^{-\tau/T_{1a}})} \frac{P}{\frac{R}{\lambda}}$$

Here, $\boldsymbol{P}$ is the mean perfusion-weighted image signal, $\boldsymbol{R}$ is the reference image signal, $\boldsymbol{\varepsilon}$ is the combined efficiency of labelling and background suppression (~65%), $\boldsymbol{\tau}$ is the label duration (1525 ms), $\boldsymbol{w}$ is the post-labelling delay (2025 ms), and $\boldsymbol{T_{1a}}$ is the $\boldsymbol{T_1}$ of arterial water.

**ASL Image Processing**

All MRI data underwent initial quality checks to identify artifacts. Complete ASL data sets were available for 25 participants, after data from one participant was excluded due to image artifact.

A multi-step pipeline was employed for the spatial normalisation of the CBF maps to MNI152 space that included: 1) co-registration of the proton density image from each sequence to the participant's T1-W image after resetting the origin of both images to the anterior commissure. The transformation matrix of this co-registration step was then applied to the CBF maps, to transform these to the T1-W image space; 2) unified segmentation of the T1-W image to generate a binary mask including only brain tissues; 3) elimination of extra-cerebral signal from the CBF maps, by multiplication of the "brain only" binary mask obtained in step 2) with each of the co-registered CBF maps; 4) normalisation of masked CBF maps (four per participant) to MNI152 space using the normalisation parameters obtained from the T1-W image segmentation in step 2). Finally, the normalised CBF maps were spatially smoothed using a 6-mm Gaussian smoothing kernel. All of these steps were implemented using the ASAP (Automatic Software for ASL processing) toolbox (version 2.0) (1) framework using the statistical parametric mapping (SPM12) software (2) run in Matlab 2021b.

**Analysis of ASL data**

***Global CBF***

To investigate the effects of ketamine and pretreatment condition on global CBF, we first extracted mean global CBF values with an explicit binary mask for grey matter using MarsBaR (3). The binary mask was derived from a standard T1-based probabilistic map of grey-matter distribution by thresholding all voxels with a probability > 0.20. We tested for the main effects of ketamine or pretreatment condition and for the ketamine-by-condition interaction on global grey-matter CBF signal using a linear mixed effects model with participant ID included as a random intercept in the model.

*Region of interest analyses*

We tested the effects of ketamine, pretreatment condition and ketamine × pretreatment condition interactions on mean rCBF values extracted using the *MarsBaR* toolbox from nine regions-of-interest (ROIs), corresponding to anatomical areas shown to be affected by ketamine administration (4–7) (**Figure S1**). Left and right sgACC ROIs, encompassing Brodmann area 25 (MNI coordinates ± 4, 21, − 8) were chosen as they have been used in several previous studies in MDD patients and clearly mapped on the anatomical region of the sgACC during visual inspection (8–10). A more rostral sgACC ROI, encompassing Brodmann area 32 (MNI coordinates: -1, 34, -5), and left and right dACC ROIs (MNI coordinates: ± 9, 39, 15), were created based on a meta-analysis examining structural and functional neural correlates of rumination in depressed patients (11). A pgACC ROI was created based on previous work examining acute effects of ketamine (12) (MNI coordinates: 0, 42, 2). The sgACC, pgACC and dACC ROIs were all defined as 5 mm radius spheres. A bilateral anterior insula ROI was derived from automated meta-analytic data on http://neurosynth.org/ using the 'salience' term (uniformity tests). This meta-analytic map was thresholded at an appropriate level ($Z = 10$) to achieve anatomically plausible regions, and the anterior insula clusters were isolated and binarized for use as an image mask. Finally, subcortical ROIs (thalamus and hippocampus) were defined using FSL's Harvard–Oxford Subcortical Structural Atlas probabilistic maps (13) and thresholded to generate binary masks. Thalamus was thresholded at 80% to maximise anatomical specificity (minimising extra-thalamic voxels) (14), and hippocampus at 20% to maintain coverage of anatomically variable structure (15). ROI masks were visually inspected to confirm appropriate coverage.

For the ROI analyses linear mixed effects models were used, including ketamine, pretreatment condition and ketamine × pretreatment condition as fixed effects, participants as a random effect and global grey-matter CBF as a nuisance covariate. All analyses were implemented using R software (version 4.2.1) with the nlme package for linear mixed-effects modelling (16). The Benjamini-Hochberg procedure was used to maintain the false discovery rate at a 5% significance level ($\alpha = 0.05$) (17). Separate sensitivity linear mixed-effects models were conducted to assess the influence of potential confounders on rCBF. Relative to the primary model, we (i) added washout duration (visit interval) as a covariate and (ii) ketamine × condition × order interactions to evaluate whether the pre–post ketamine effect under naltrexone versus placebo differed by treatment order (Placebo-Naltrexone vs Naltrexone-Placebo) (**see Supplementary Material: Sensitivity Analyses**).

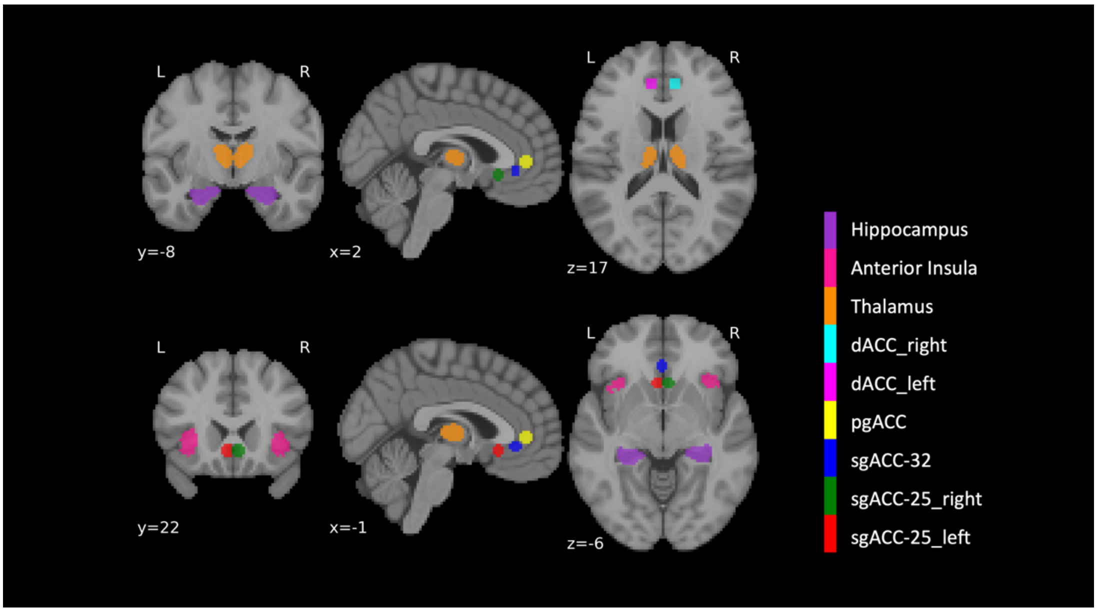


**eFigure1: Visualisation of the nine regions of interest overlaid on MNI 152 T1-weighted template image.** MNI (x, y, z) coordinates are displayed.

***Whole brain voxel-wise analyses***

Processed whole-brain CBF images were analysed using a voxel-wise flexible factorial design in SPM (2) to investigate ketamine, pretreatment condition and ketamine-by- pretreatment condition effects on voxel-wise CBF. Alongside the overall main ketamine effect, the effect of ketamine was also assessed for each pretreatment condition separately. Analyses were restricted to grey matter tissue using a binarized grey matter mask and to account for sex and age-related variability in CBF (18), sex and age were included as nuisance variables. Finally, global-grey matter CBF was also included as a nuisance covariate (i.e., global normalisation). Results without global normalisation are presented as supplementary. Statistical maps were thresholded at $p < 0.05$, voxel-wise (peak-level) FWE-corrected.

***Associations between clinical and subjective measures and regional CBF***

We examined the relationship between baseline rCBF (pre-ketamine infusion) and day 1 post-infusion clinical measure change scores (MADRS, QIDS-SR, SHAPS, TEPS-A and TEPS-C) that have previously been reported (19). We estimated the partial Pearson's correlation coefficients between mean baseline (pre-ketamine infusion) rCBF values extracted from each of the nine pre-defined ROIs and the day 1 clinical measure change scores (day 1 scores minus pre-infusion baseline scores), adjusting for global CBF. Next, we examined the relationship between rCBF changes during the ketamine infusion and day 1 post-infusion clinical measure change scores. We first calculated relative rCBF values as the ratio of regional (absolute) CBF to global CBF for each of the nine ROIs for the pre-ketamine and ketamine-infusion scans. We then estimated the partial Pearson's correlation coefficients between the relative rCBF values for each ROI from the ketamine infusion scan and the day 1 clinical measure change scores, adjusting for relative rCBF values from the pre-ketamine infusion scan. We similarly explored the relationship between baseline rCBF and changes in relative rCBF during the ketamine infusion and acute subjective measure scores (CADSS and PSI subscale scores). For these correlation analyses we applied a Bonferroni correction according to the number of ROIs (0.05/9), setting a threshold for statistical significance at $p < 0.0056$.

We estimated these correlations separately for placebo and naltrexone pretreatment conditions. For completeness, if any significant correlations were identified we then statistically compared the partial correlation coefficients for placebo and naltrexone conditions using the Steiger Z-transform test (for dependent correlations) (20), implemented using r.test from the 'psych' library in R (21).

***Correlations between ΔCBF and receptor density profiles***

We assessed the spatial correlation between ketamine induced rCBF changes and receptor distributions alongside the effect of naltrexone. **eFigure S2** provides an overview of the approach used for this analysis. First, we examined the direct impact of ketamine in the placebo pretreatment condition, avoiding naltrexone contamination (*Placebo: Ketamine > Pre-ketamine*). Second, we assessed the interaction between ketamine and naltrexone pretreatment *(Interaction: Ketamine x Naltrexone)*. Unthresholded contrast images without global normalisation, with a whole brain mask applied, were first extracted and then parcellated to 422 regions according to a combined parcellation atlas (400 cortical regions from the Schaefer atlas (22) and 22 subcortical regions (23)).

We obtained volumetric PET images, indicative of receptor densities, from public sources (24) for various receptors. Specifically, we studied MOR, KOR, NMDA, mGluR5, $GABA_A$, and $GABA_A\alpha5$, which utilise the following radiotracers: [$^{11}$C]Carfentanil (25), [$^{11}$C]LY2795050 (26), [$^{18}$F]GE-179 (27), [$^{11}$C]ABP688 (28,29), [$^{11}$C]Flumazenil (30) and [$^{11}$C]Ro15-4513 (31) respectively. These images are an estimate proportional to receptor densities that reflect binding potential and tracer distribution volume, which we refer to simply as density. These receptor maps were chosen based on our theory that both the glutamatergic and opioid systems play a role in ketamine's effects and that naltrexone could potentially influence these effects through these systems. Additionally, we focussed on the GABAergic system, as it may facilitate the interaction between the opioid and glutamatergic systems.

Considering potential interactions between the opioid and dopaminergic and serotonergic systems in ketamine's antidepressant mechanisms (32–34), we further extended our analysis to include a range of dopaminergic ($D_1$ and $D_2$) and ($5HT_{1A}$, $5HT_{1B}$, $5HT_{2A}$, $5HT_4$, $5HT_6$, and 5HTT) PET maps using the following tracers: [$^{11}$C]SCH23390 (35) , [$^{11}$C]FLB-457 (36,37), [$^{11}$C]WAY-100635 (38), [$^{11}$C]P943 (39), [$^{11}$C]CIMBI-36 (40), [$^{11}$C]SB207145 (40), [$^{11}$C]GSK215083 (41), [$^{11}$C]DASB (40).

PET images were all registered to the MNI-ICBM 152 non-linear 2009 (version c, asymmetric) template and then parcellated to 422 regions according to the combined cortical and subcortical atlas also used to parcellate the CBF contrast maps. For mGluR5, $D_2$ and $5HT_{1B}$ there were multiple mean images using the same tracer that were highly correlated to one another. We employed a weighted average method to integrate these images, consistent with approaches previously established in the literature (42).

Spearman's correlation coefficients were calculated to examine the spatial relationship between the ΔCBF contrast profiles and each receptor density profile. These observed values were compared to a null distribution generated from 10,000 spatial-autocorrelation (SA)-preserving surrogate maps for each ΔCBF contrast profile, using the BrainSMASH (brain surrogate maps with autocorrelated spatial heterogeneity) python toolbox so as to account for patterns of spatial autocorrelation (43). Given the exploratory nature of this analysis, we reported all correlations that withstood the SA-correction ($p < 0.05$) but also highlight any correlations surviving an additional FDR correction.

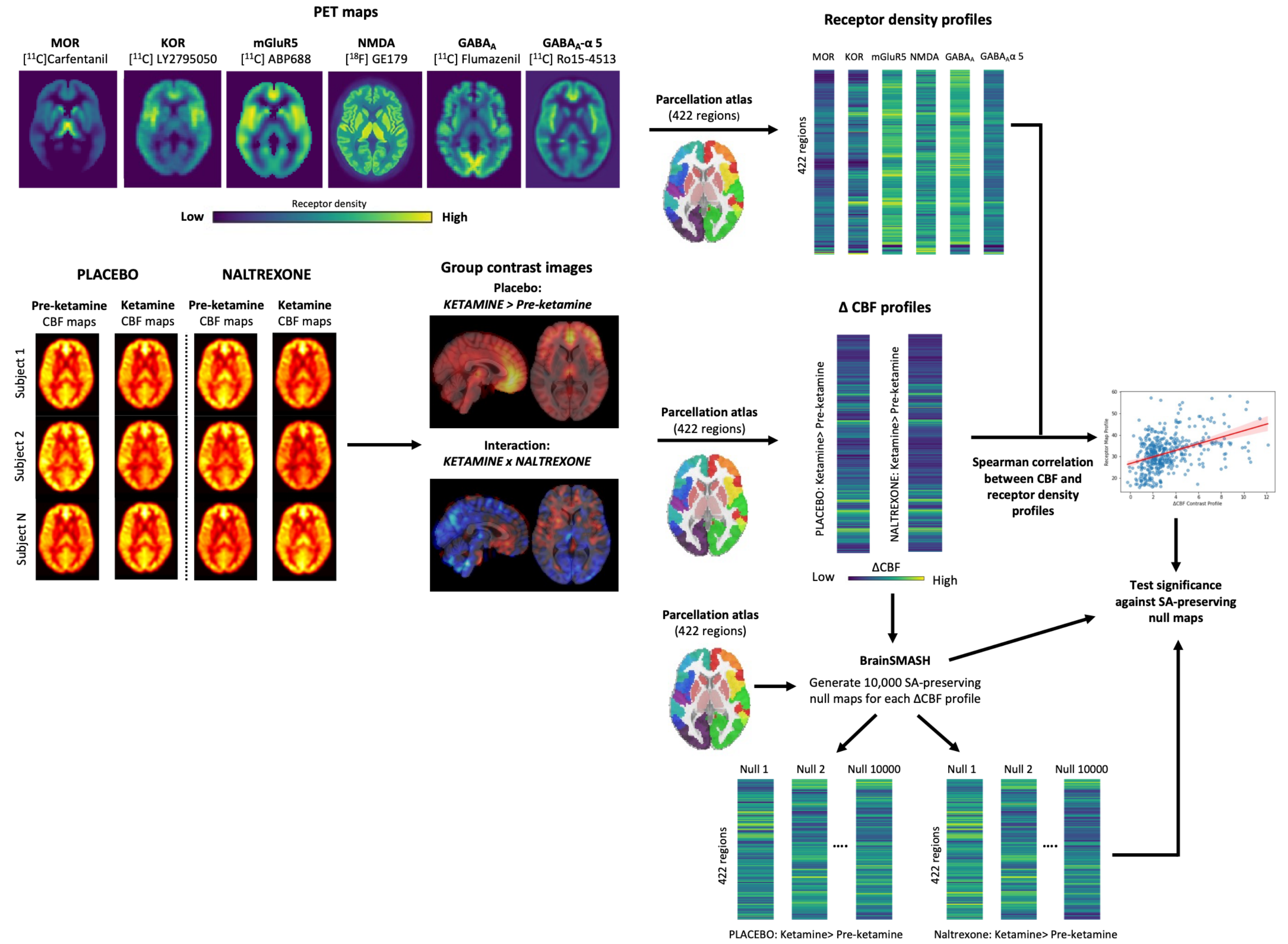


**eFigure 2: Overview of approach to examine correlations between ΔCBF and receptor density profiles.** 1) PET images registered to MNI-152 template and parcellated into 422 regions according to parcellation atlas. In addition to the MOR, KOR mGluR5, NMDA, $GABA_A$ and $GABA_A\alpha5$ PET maps shown in the figure, supplementary analyses also included $D_1$ $D_2$, $5HT_{1A}$, $5HT_{1B}$, $5HT_{2A}$, $5HT_4$, $5HT_6$, and 5HTT PET maps; 2) Unthresholded contrast images generated for a) Placebo: *'Ketamine > Pre-ketamine'* and b) the interaction effect: '*Ketamine x Naltrexone'*, which were each parcellated into 422 regions according to parcellation atlas; 3) For each ΔCBF contrast profile, a null distribution was generated from 10,000 spatial-autocorrelation (SA)-preserving surrogates, using the BrainSMASH toolbox to account for patterns of spatial autocorrelation**; 4)** Spearman's correlation coefficients were calculated to examine the spatial relationship between the ΔCBF contrast profiles and each receptor density profile, adjusting significance according to the 10,000 SA-preserving null maps.

### Supplementary Results

#### Global CBF

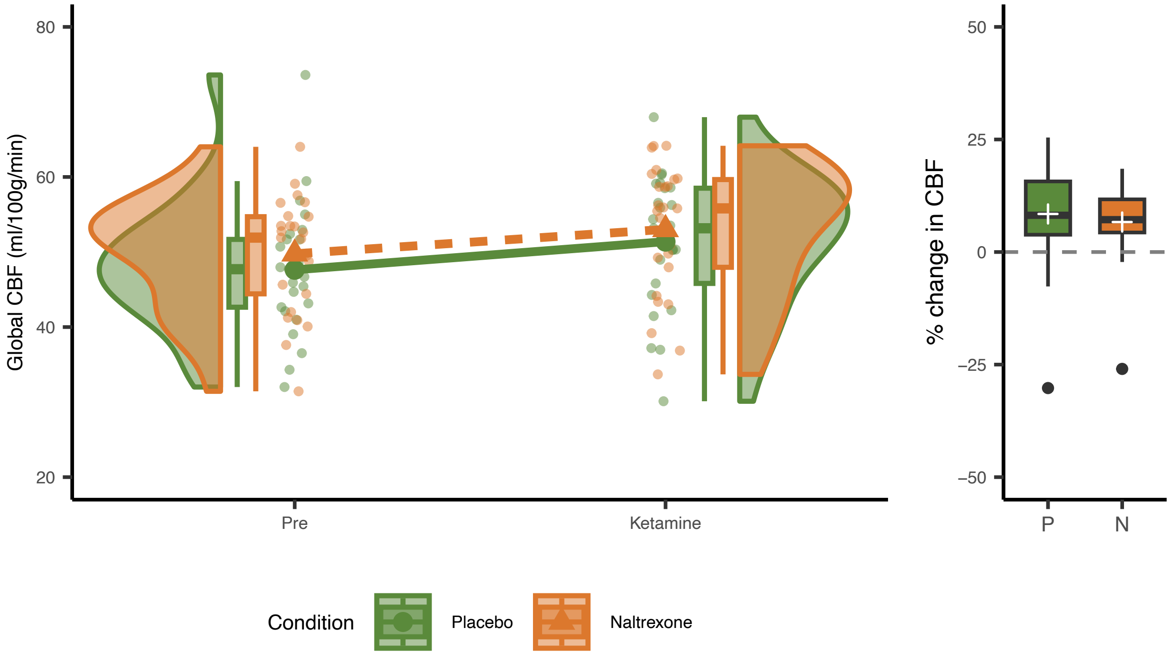


**eFigure 3: Global CBF values.** Global CBF values are shown for the Pre (pre-infusion) and Ketamine (during infusion) scans under both placebo and naltrexone pretreatment conditions (left panel). The thick lines represent the mean values for each condition. The percentage change in global CBF from Pre to Ketamine is displayed for each condition—placebo (P) and naltrexone (N)—in the boxplot (right panel). Box plot elements: Box spans the interquartile range (IQR) (25th–75th percentile), central line is the median (50th percentile), "+" marks the mean, whiskers extend to the lowest and highest values within 1.5 × IQR of the box, and values beyond that range are shown individually. Each plot shows data from participants with complete ASL data sets (n = 25).

**Region of interest analyses**

**eTable 2: Regional CBF linear mixed-effects model results for selected regions of interest.**

| | KETAMINE | | | CONDITION | | | KETAMINE x CONDITION | | |
|---|---|---|---|---|---|---|---|---|---|
| ROI | F Value | Uncorrected p-value | Adjusted p-value | F Value | Uncorrected p-value | Adjusted p-value | F Value | Uncorrected p-value | Adjusted p-value |
| **sgACC-25_L** | 80.24 | < 0.001 | < 0.001 | 20.75 | < 0.001 | < 0.001 | 1.90 | 0.172 | 0.222 |
| **sgACC-25_R** | 88.71 | < 0.001 | < 0.001 | 19.68 | < 0.001 | < 0.001 | 3.13 | 0.081 | 0.115 |
| **sgACC-32** | 115.10 | < 0.001 | < 0.001 | 9.97 | 0.002 | 0.004 | 1.22 | 0.274 | 0.314 |
| **pgACC** | 270.04 | < 0.001 | < 0.001 | 23.61 | < 0.001 | < 0.001 | 1.19 | 0.279 | 0.314 |
| **dACC_L** | 55.24 | < 0.001 | < 0.001 | 10.44 | 0.002 | 0.003 | 0.40 | 0.529 | 0.550 |
| **dACC_R** | 130.18 | < 0.001 | < 0.001 | 28.20 | < 0.001 | < 0.001 | 0.69 | 0.408 | 0.441 |
| **Thalamus** | 81.39 | < 0.001 | < 0.001 | 14.18 | < 0.001 | 0.001 | 1.00 | 0.260 | 0.314 |
| **Insula** | 337.02 | < 0.001 | < 0.001 | 43.00 | < 0.001 | < 0.001 | 2.75 | 0.102 | 0.137 |
| **Hippocampus** | 28.17 | < 0.001 | < 0.001 | 16.88 | < 0.001 | < 0.001 | 0.04 | 0.838 | 0.838 |

**KETAMINE**- main effect of ketamine; **CONDITION**- effect of pretreatment condition; **KETAMINE x CONDITION**- Ketamine by pretreatment condition interaction effect; **Adjusted p-value-** corrected for false discovery rate using Benjamini–Hochberg procedure across all ROI tests reported (9 ROIs × 3 planned contrasts; 27 tests).

**Sensitivity analyses**

Adding washout duration as a covariate did not materially change the results shown in eTable2. The main effects of ketamine (pre–post ketamine) and condition (naltrexone vs placebo) remained significant across ROIs, and there were no significant ketamine × condition interactions.

To assess whether the pre–post ketamine effect under naltrexone versus placebo differed by treatment order (Placebo-Naltrexone vs Naltrexone-Placebo), we tested the ketamine × condition × order interaction. No ROIs showed a significant ketamine × condition × order interaction, indicating that there was no order-dependent naltrexone-versus-placebo difference in the ketamine pre–post rCBF change. We did observe significant ketamine × order interactions for sgACC_32 ($p = 0.013$), pgACC ($p = 0.008$), dACC_L ($p = 0.001$), and insula ($p = 0.035$), suggesting that the overall pre–post ketamine regional CBF change differed by order in these ROIs, with a larger pre–post change in the Placebo-Naltrexone sequence (order 1) than in the Naltrexone-Placebo sequence (order 2). However, because there was no evidence that order modified the naltrexone-versus-placebo difference (ketamine × condition × order non-significant), these findings do not change the interpretation of the primary results.

**Whole-brain voxel-wise analysis**

**eTable : Brain regions demonstrating significant increases in rCBF for main effect of ketamine administration (with global normalisation).**

| Statistical Test | Regions | Cluster-level | | Peak-level (Primary inference) | | MNI coordinates | | |
|---|---|---|---|---|---|---|---|---|
| | | $p_{FWE}$ | $k_E$ | $p_{FWE}$ | T | x | y | z |
| **Ketamine main effect**<br><br>(Ketamine > Pre-ketamine) | sgACC, pgACC, dACC, anterior PFC, bilateral insula, inferior frontal gyri (pars orbitalis), orbitofrontal cortex | <0.001 | 5749 | <0.001 | 10.29 | 48 | 40 | -10 |
| | | | | *<0.001* | *9.70* | *14* | *16* | *-26* |
| | | | | *<0.001* | *9.07* | *38* | *40* | *-20* |
| | | 0.028 | 2 | 0.038 | 5.17 | -32 | 54 | -4 |
| | | 0.028 | 2 | 0.039 | 5.17 | 4 | 38 | 40 |
| | | 0.028 | 2 | 0.044 | 5.13 | 52 | 22 | 18 |

SPM12 whole-brain results. For each contrast, we report cluster extent (kE), cluster-level FWE-corrected p-values (cluster $p_{FWE}$), and voxel-wise (peak-level) FWE-corrected p-values (peak $p_{FWE}$) with associated T-values. Clusters were formed at the voxel-wise FWE height threshold ($p_{FWE} < 0.05$; $T > 5.09$) with no extent threshold ($k = 0$). MNI coordinates (x, y, z) correspond to the peak voxel of each cluster; greyed rows indicate additional local maxima (sub-peaks) within the same cluster. Primary inference is based on peak-level (voxel-wise) pFWE.

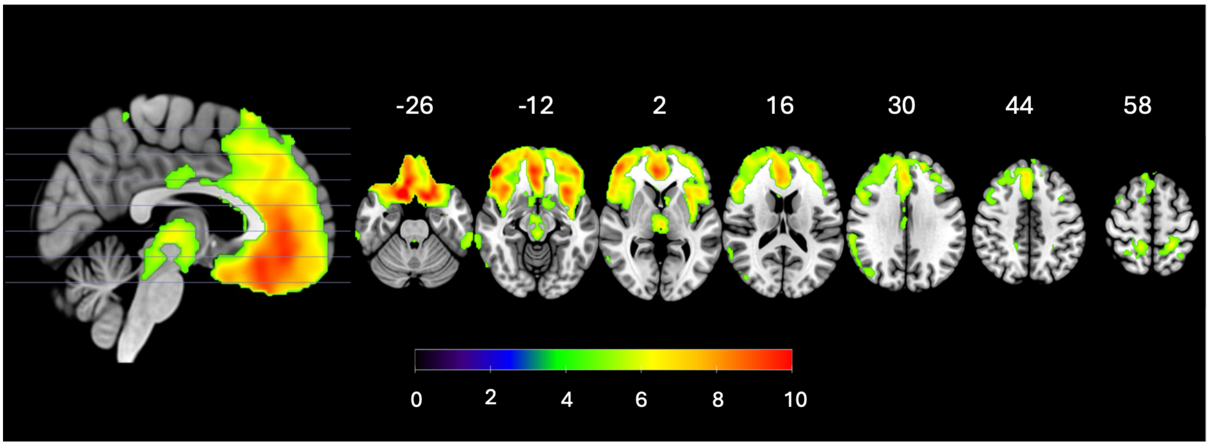


**eFigure 4: The main effect of ketamine on CBF for the whole-brain voxel-wise analysis (without global normalisation).** Results overlaid on the SPM152 T1 template distributed with MRIcroGL. The MNI Z axis is shown at the top of the figure. Statistical maps were thresholded at $p < 0.05$, voxel-wise (peak-level) FWE-corrected. The colour bar shows the t statistic.

**ΔCBF and receptor density profile associations**

**eTable 4: Associations between ΔCBF and receptor density profiles.**

| | **Placebo: Ketamine > Pre-ketamine** | | | | | **Interaction: Ketamine x Naltrexone** | | | | |
|---|---|---|---|---|---|---|---|---|---|---|
| **Receptor** | **Spearman r** | **p-value (unadjusted)** | **$p_{SA\text{-}corr}$** | **Null SD** | **Z-score** | **Spearman r** | **p-value (unadjusted)** | **$p_{SA\text{-}corr}$** | **Null SD** | **Z-score** |
| **MOR** | **0.512** | **<0.0001** | **0.0209 *** | **0.239** | **2.164** | **0.539** | **<0.0001** | **0.0194 *** | **0.256** | **2.106** |
| **KOR** | 0.300 | <0.0001 | 0.1386 | 0.198 | 1.531 | 0.388 | <0.0001 | 0.0550 | 0.210 | 1.855 |
| **mGluR5** | **0.348** | **<0.0001** | **0.0038 **** | **0.130** | **2.686** | **0.257** | **<0.0001** | **0.0387 *** | **0.128** | **2.025** |
| **NMDA** | 0.102 | 0.0371 | 0.3865 | 0.112 | 0.920 | 0.158 | 0.0012 | 0.1747 | 0.114 | 1.381 |
| **GABAA** | 0.034 | 0.4848 | 0.7987 | 0.121 | 0.265 | −0.158 | 0.0011 | 0.2210 | 0.124 | −1.267 |
| **GABAAα5** | 0.318 | <0.0001 | 0.1196 | 0.202 | 1.589 | **0.520** | **<0.0001** | **0.0021 **** | **0.213** | **2.454** |

For each CBF contrast (‘*Placebo: Ketamine > Pre-ketamine*’ and ‘*Interaction: Ketamine x Naltrexone*’) the correlation coefficient with each receptor map is shown, alongside the unadjusted p-value and the p-value after spatial autocorrelation adjustment ($p_{SA\text{-}corr}$). Null standard deviations (SD) and corresponding z-scores are also shown. Bold values are correlations that remained statistically significant after the spatial autocorrelation adjustment (*: $p_{SA\text{-}corr} < 0.05$). (**: Indicates statistical significance surviving additional Benjamini-Hochberg FDR correction)

**ΔCBF, dopaminergic and serotonergic receptor density profile associations**

These exploratory analyses revealed significant correlations between ketamine-induced ΔCBF and the $5HT_{1B}$ profile ($r = 0.369$, $p_{SA\text{-}corr} = 0.0069$) (**Table S5**). For the ketamine x naltrexone interaction CBF contrast, there were correlations with $D_1$, $D_2$, $5HT_{1A}$, $5HT_4$ and 5HTT receptor density profiles that also remained significant after spatial autocorrelation adjustment.

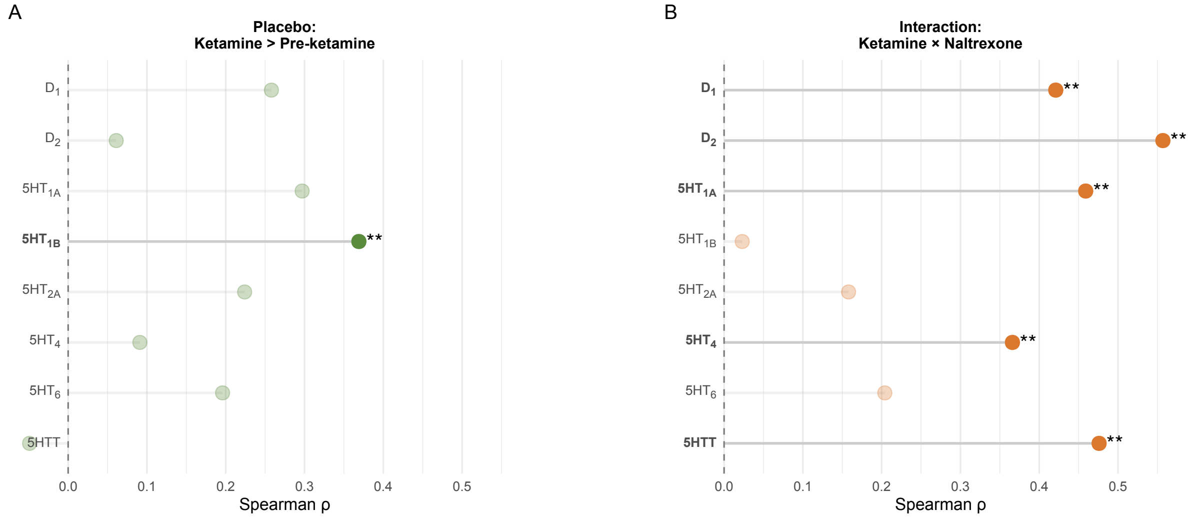


**eFigure 5: Associations between ΔCBF and dopaminergic and serotonergic receptor density profiles.** (A) Main effect of ketamine under placebo (Ketamine > Pre-Ketamine) with Spearman correlations against each receptor map. (B) Ketamine × Naltrexone interaction effect on CBF with Spearman correlations against each receptor map. Asterisks denote correlations significant after spatial-autocorrelation correction (* pSA-corr < 0.05) and after additional Benjamini–Hochberg FDR correction (** FDR-corr < 0.05).

**eTable 5: Associations between ΔCBF and dopaminergic and serotonergic receptor density profiles.**

| | Placebo: Ketamine > Pre-ketamine | | | | | Interaction: Ketamine x Naltrexone | | | | |
|---|---|---|---|---|---|---|---|---|---|---|
| Receptor | Spearman r | p-value (unadjusted) | $p_{SA\text{-}corr}$ | Null SD | Z-score | Spearman r | p-value (unadjusted) | $p_{SA\text{-}corr}$ | Null SD | Z-score |
| D1 | 0.258 | <0.0001 | 0.1636 | 0.180 | 1.454 | **0.421** | **<0.0001** | **0.0123 **** | **0.188** | **2.239** |
| D2 | 0.061 | 0.2110 | 0.7992 | 0.215 | 0.302 | **0.557** | **<0.0001** | **0.0013 **** | **0.223** | **2.487** |
| 5HT1A | 0.297 | <0.0001 | 0.0936 | 0.178 | 1.685 | **0.459** | **<0.0001** | **0.0021 **** | **0.185** | **2.487** |
| 5HT1B | **0.369** | **<0.0001** | **0.0069 **** | **0.146** | **2.528** | 0.023 | 0.6434 | 0.8920 | 0.153 | 0.159 |
| 5HT2A | 0.224 | <0.0001 | 0.1053 | 0.135 | 1.653 | 0.158 | 0.0012 | 0.2757 | 0.138 | 1.156 |
| 5HT4 | 0.091 | 0.0630 | 0.6159 | 0.164 | 0.567 | **0.366** | **<0.0001** | **0.0163 **** | **0.170** | **2.154** |
| 5HT6 | 0.196 | <0.0001 | 0.1157 | 0.122 | 1.606 | 0.204 | <0.0001 | 0.0874 | 0.120 | 1.704 |
| 5HTT | −0.049 | 0.3190 | 0.8218 | 0.196 | −0.231 | **0.476** | **<0.0001** | **0.0092 **** | **0.204** | **2.326** |

For each CBF contrast (‘*Placebo: Ketamine > Pre-ketamine’* and ‘*Interaction: Ketamine x Naltrexone’*) the correlation coefficient with each receptor map is shown, alongside the unadjusted p-value and the p-value after spatial autocorrelation adjustment ($p_{SA\text{-}corr}$). Null standard deviations (SD) and corresponding z-scores are also shown. Bold values are correlations that remained statistically significant after the spatial autocorrelation adjustment (*$p_{SA\text{-}corr}<0.05$). (**: Indicates statistical significance surviving additional Benjamini-Hochberg FDR correction)

### References


1. Mato Abad V, García-Polo P, O'Daly O, Hernández-Tamames JA, Zelaya F. ASAP (Automatic Software for ASL Processing): A toolbox for processing Arterial Spin Labeling images. Magnetic Resonance Imaging. 2016 Apr 1;34(3):334–44. doi:10.1016/j.mri.2015.11.002

2. SPM12. SPM12 [Internet]. http://fil.ion.ucl.ac.uk/spm/. Available from: http://fil.ion.ucl.ac.uk/spm/.

3. Brett M, Anton JL, Valabregue R, Poline JB. Region of interest analysis using an SPM toolbox. In. Sendai, Japan; 2002.

4. Alexander L, Jelen LA, Mehta MA, Young AH. The anterior cingulate cortex as a key locus of ketamine's antidepressant action. Neurosci Biobehav Rev. 2021 Aug;127:531–54. doi:10.1016/j.neubiorev.2021.05.003 PubMed PMID: 33984391.

5. Bojesen KB, Andersen KA, Rasmussen SN, Baandrup L, Madsen LM, Glenthoj BY, et al. Glutamate Levels and Resting Cerebral Blood Flow in Anterior Cingulate Cortex Are Associated at Rest and Immediately Following Infusion of S-Ketamine in Healthy Volunteers. Front Psychiatry. 2018/02/23 edn. 2018;9:22. Located at: 29467681. doi:10.3389/fpsyt.2018.00022

6. Bryant JE, Frölich M, Tran S, Reid MA, Lahti AC, Kraguljac NV. Ketamine induced changes in regional cerebral blood flow, interregional connectivity patterns, and glutamate metabolism. Journal of Psychiatric Research. 2019 Oct 1;117:108–15. doi:10.1016/j.jpsychires.2019.07.008

7. Pollak TA, De Simoni S, Barimani B, Zelaya FO, Stone JM, Mehta MA. Phenomenologically distinct psychotomimetic effects of ketamine are associated with cerebral blood flow changes in functionally relevant cerebral foci: a continuous arterial spin labelling study. Psychopharmacology (Berl). 2015/10/07 edn. 2015 Dec;232(24):4515–24. Located at: 26438425. doi:10.1007/s00213-015-4078-8

8. Berwian IM, Wenzel JG, Kuehn L, Schnuerer I, Kasper L, Veer IM, et al. The relationship between resting-state functional connectivity, antidepressant discontinuation and depression relapse. Sci Rep. 2020 Dec 18;10:22346. doi:10.1038/s41598-020-79170-9 PubMed PMID: 33339879; PubMed Central PMCID: PMC7749105.

9. Bhaumik R, Jenkins LM, Gowins JR, Jacobs RH, Barba A, Bhaumik DK, et al. Multivariate pattern analysis strategies in detection of remitted major depressive disorder using resting state functional connectivity. NeuroImage: Clinical. 2017 Jan 1;16:390–8. doi:10.1016/j.nicl.2016.02.018

10. Jenkins LM, Stange JP, Barba A, DelDonno SR, Kling LR, Briceño EM, et al. Integrated cross-network connectivity of amygdala, insula, and subgenual cingulate associated with facial emotion perception in healthy controls and remitted major depressive disorder. Cogn Affect Behav Neurosci. 2017 Dec 1;17(6):1242–54. doi:10.3758/s13415-017-0547-3

11. Kühn S, Vanderhasselt MA, De Raedt R, Gallinat J. Why ruminators won't stop: The structural and resting state correlates of rumination and its relation to depression. Journal of Affective Disorders. 2012 Dec 10;141(2):352–60. doi:10.1016/j.jad.2012.03.024

12. Gärtner M, Weigand A, Scheidegger M, Lehmann M, Wyss PO, Wunder A, et al. Acute effects of ketamine on the pregenual anterior cingulate: linking spontaneous activation, functional connectivity, and glutamate metabolism. Eur Arch Psychiatry Clin Neurosci. 2022 Jun;272(4):703–14. doi:10.1007/s00406-021-01377-2

13. Jenkinson M, Beckmann CF, Behrens TEJ, Woolrich MW, Smith SM. FSL. Neuroimage. 2012 Aug 15;62(2):782–90. doi:10.1016/j.neuroimage.2011.09.015 PubMed PMID: 21979382.

14. Abram SV, Roach BJ, Fryer SL, Calhoun VD, Preda A, van Erp TGM, et al. Validation of ketamine as a pharmacological model of thalamic dysconnectivity across the illness course of schizophrenia. Mol Psychiatry. 2022 May;27(5):2448–56. doi:10.1038/s41380-022-01502-0

15. Lambert HK, Sheridan MA, Sambrook KA, Rosen ML, Askren MK, McLaughlin KA. Hippocampal Contribution to Context Encoding across Development Is Disrupted following Early-Life Adversity. J Neurosci. 2017 Feb 15;37(7):1925–34. doi:10.1523/JNEUROSCI.2618-16.2017 PubMed PMID: 28093475.

16. Pinheiro J, Bates D. nlme: Linear and Nonlinear Mixed Effects Models [Internet]. 2022. Available from: https://cran.r-project.org/web/packages/nlme/nlme.pdf

17. Feser WJ, Fingerlin TE, Strand MJ, Glueck DH. CALCULATING AVERAGE POWER FOR THE BENJAMINI-HOCHBERG PROCEDURE. J Stat Theory Appl. 2009;8(3):325–52. PubMed PMID: 27818616; PubMed Central PMCID: PMC5095931.

18. Alisch JSR, Khattar N, Kim RW, Cortina LE, Rejimon AC, Qian W, et al. Sex and age-related differences in cerebral blood flow investigated using pseudo-continuous arterial spin labeling magnetic resonance imaging. Aging (Albany NY). 2021 Feb 17;13(4):4911–25. doi:10.18632/aging.202673 PubMed PMID: 33596183; PubMed Central PMCID: PMC7950235.

19. Jelen LA, Lythgoe DJ, Stone JM, Young AH, Mehta MA. Effect of naltrexone pretreatment on ketamine-induced glutamatergic activity and symptoms of depression: a randomized crossover study. Nat Med. 2025 Jul 24;1–9. doi:10.1038/s41591-025-03800-w

20. Steiger JH. Testing Pattern Hypotheses On Correlation Matrices: Alternative Statistics And Some Empirical Results. Multivariate Behavioral Research. 1980 Jul 1;15(3):335–52. doi:10.1207/s15327906mbr1503_7 PubMed PMID: 26794186.

21. Revelle W. psych: Procedures for Psychological, Psychometric, and Personality Research [Internet]. 2022. Available from: https://cran.r-project.org/web/packages/psych/index.html

22. Schaefer A, Kong R, Gordon EM, Laumann TO, Zuo XN, Holmes AJ, et al. Local-Global Parcellation of the Human Cerebral Cortex from Intrinsic Functional Connectivity MRI. Cereb Cortex. 2018 Sep 1;28(9):3095–114. doi:10.1093/cercor/bhx179 PubMed PMID: 28981612; PubMed Central PMCID: PMC6095216.

23. Pauli WM, Nili AN, Tyszka JM. A high-resolution probabilistic in vivo atlas of human subcortical brain nuclei. Sci Data. 2018 Apr 17;5(1):1. doi:10.1038/sdata.2018.63

24. Markello RD, Hansen JY, Liu ZQ, Bazinet V, Shafiei G, Suárez LE, et al. neuromaps: structural and functional interpretation of brain maps. Nat Methods. 2022 Nov;19(11):11. doi:10.1038/s41592-022-01625-w

25. Kantonen T, Karjalainen T, Isojärvi J, Nuutila P, Tuisku J, Rinne J, et al. Interindividual variability and lateralization of μ-opioid receptors in the human brain. NeuroImage. 2020 Aug 15;217:116922. doi:10.1016/j.neuroimage.2020.116922

26. Naganawa M, Zheng MQ, Nabulsi N, Tomasi G, Henry S, Lin SF, et al. Kinetic modeling of 11C-LY2795050, a novel antagonist radiotracer for PET imaging of the kappa opioid receptor in humans. J Cereb Blood Flow Metab. 2014 Nov;34(11):1818–25. doi:10.1038/jcbfm.2014.150 PubMed PMID: 25182664; PubMed Central PMCID: PMC4269759.

27. Galovic M, Erlandsson K, Fryer TD, Hong YT, Manavaki R, Sari H, et al. Validation of a combined image derived input function and venous sampling approach for the quantification of [18F]GE-179 PET binding in the brain. NeuroImage. 2021 Aug 15;237:118194. doi:10.1016/j.neuroimage.2021.118194

28. DuBois JM, Rousset OG, Rowley J, Porras-Betancourt M, Reader AJ, Labbe A, et al. Characterization of age/sex and the regional distribution of mGluR5 availability in the healthy human brain measured by high-resolution [11C]ABP688 PET. Eur J Nucl Med Mol Imaging. 2016 Jan 1;43(1):152–62. doi:10.1007/s00259-015-3167-6

29. Smart K, Cox SML, Scala SG, Tippler M, Jaworska N, Boivin M, et al. Sex differences in [11C]ABP688 binding: a positron emission tomography study of mGlu5 receptors. Eur J Nucl Med Mol Imaging. 2019 May 1;46(5):1179–83. doi:10.1007/s00259-018-4252-4

30. Nørgaard M, Beliveau V, Ganz M, Svarer C, Pinborg LH, Keller SH, et al. A high-resolution in vivo atlas of the human brain's benzodiazepine binding site of GABAA receptors. NeuroImage. 2021 May 15;232:117878. doi:10.1016/j.neuroimage.2021.117878

31. Horder J, Andersson M, Mendez MA, Singh N, Tangen Ä, Lundberg J, et al. GABAA receptor availability is not altered in adults with autism spectrum disorder or in mouse models. Science Translational Medicine. 2018 Oct 3;10(461):eaam8434. doi:10.1126/scitranslmed.aam8434

32. Jelen LA, Stone JM, Young AH, Mehta MA. The opioid system in depression. Neurosci Biobehav Rev. 2022 Sep;140:104800. doi:10.1016/j.neubiorev.2022.104800 PubMed PMID: 35914624.

33. Jelen LA, Young AH, Stone JM. Ketamine: A tale of two enantiomers. J Psychopharmacol. 2020/11/07 edn. 2021 Feb;35(2):109–23. Located at: 33155503. doi:10.1177/0269881120959644

34. Kokkinou M, Ashok AH, Howes OD. The effects of ketamine on dopaminergic function: meta-analysis and review of the implications for neuropsychiatric disorders. Mol Psychiatry. 2017/10/04 edn. 2018 Jan;23(1):59–69. Located at: 28972576. doi:10.1038/mp.2017.190

35. Kaller S, Rullmann M, Patt M, Becker GA, Luthardt J, Girbardt J, et al. Test-retest measurements of dopamine D1-type receptors using simultaneous PET/MRI imaging. Eur J Nucl Med Mol Imaging. 2017 Jun;44(6):1025–32. doi:10.1007/s00259-017-3645-0 PubMed PMID: 28197685.

36. Sandiego CM, Gallezot JD, Lim K, Ropchan J, Lin S fei, Gao H, et al. Reference region modeling approaches for amphetamine challenge studies with [11C]FLB 457 and PET. J Cereb Blood Flow Metab. 2015 Mar 31;35(4):623–9. doi:10.1038/jcbfm.2014.237 PubMed PMID: 25564239; PubMed Central PMCID: PMC4420880.

37. Smith CT, Crawford JL, Dang LC, Seaman KL, San Juan MD, Vijay A, et al. Partial-volume correction increases estimated dopamine D2-like receptor binding potential and reduces adult age differences. J Cereb Blood Flow Metab. 2019 May;39(5):822–33. doi:10.1177/0271678X17737693 PubMed PMID: 29090626; PubMed Central PMCID: PMC6498753.

38. Savli M, Bauer A, Mitterhauser M, Ding YS, Hahn A, Kroll T, et al. Normative database of the serotonergic system in healthy subjects using multi-tracer PET. NeuroImage. 2012 Oct 15;63(1):447–59. doi:10.1016/j.neuroimage.2012.07.001

39. Gallezot JD, Nabulsi N, Neumeister A, Planeta-Wilson B, Williams WA, Singhal T, et al. Kinetic Modeling of the Serotonin 5-HT1B Receptor Radioligand [11C]P943 in Humans. J Cereb Blood Flow Metab. 2010 Jan 1;30(1):196–210. doi:10.1038/jcbfm.2009.195

40. Beliveau V, Ganz M, Feng L, Ozenne B, Højgaard L, Fisher PM, et al. A High-Resolution In Vivo Atlas of the Human Brain's Serotonin System. J Neurosci. 2017 Jan 4;37(1):120–8. doi:10.1523/JNEUROSCI.2830-16.2016 PubMed PMID: 28053035.

41. Radhakrishnan R, Nabulsi N, Gaiser E, Gallezot JD, Henry S, Planeta B, et al. Age-Related Change in 5-HT6 Receptor Availability in Healthy Male Volunteers Measured with 11C-GSK215083 PET. J Nucl Med. 2018 Sep;59(9):1445–50. doi:10.2967/jnumed.117.206516 PubMed PMID: 29626125; PubMed Central PMCID: PMC6126437.

42. Hansen JY, Shafiei G, Markello RD, Smart K, Cox SML, Nørgaard M, et al. Mapping neurotransmitter systems to the structural and functional organization of the human neocortex. Nat Neurosci. 2022 Nov;25(11):11. doi:10.1038/s41593-022-01186-3

43. Burt JB, Helmer M, Shinn M, Anticevic A, Murray JD. Generative modeling of brain maps with spatial autocorrelation. NeuroImage. 2020 Oct 15;220:117038. doi:10.1016/j.neuroimage.2020.117038
